# Variability in Automated Sepsis Case Detection: A Systematic Analysis of Implementation Methods in Clinical Data Repositories

**DOI:** 10.64898/2026.02.27.26347259

**Authors:** Falk Meyer-Eschenbach, René Schmiedler, Johannes v. Stoephasius, Camille Zhang, Luis Kronfli, Nicolas Frey, Anatol-Fiete Näher, Jacques Ehret, Johanna Nothacker, Christof von Kalle, Severin Kohler, Elias Grünewald, Andreas Edel, Oliver Kumpf, Jon Barrenetxea, Felix Balzer

## Abstract

**Objective:** To systematically review and characterize methodological heterogeneity in sepsis case detection using the MIMIC-III and eICU-CRD databases.

**Materials and Methods:** We conducted a PRISMA-guided systematic review of PubMed and Web of Science (publication years 2016–2024). We extracted methodological details on sepsis case detection across six domains: parameter coverage, temporal windows, aggregation methods, missing-data handling, SOFA calculation, and infection detection methods. For studies with available source code, we additionally examined code structure and repository dependencies to identify methodological decisions across these domains.

**Results:** Of 396 publications screened, 64 met the inclusion criteria and 12 provided available source code. Sepsis detection rates ranged from 3.4% to 65.2% in MIMIC-III and from 9.8% to 47.9% in eICU-CRD. Substantial variability persisted among studies using identical cohort definitions within both databases (MIMIC-III: 16.9%–42.2%; eICU-CRD: 13.9%–31.4%). The overall proportion of studies reporting methodological details varied by domain: SOFA calculation (53.1%), infection detection methods (42.2%), temporal windows (37.5%), aggregation methods (26.6%) and missing-data handling (17.2%). Source code analysis identified 321 implementation decisions, revealing heterogeneity in baseline SOFA definitions (SOFA=0 vs dynamic baseline), temporal windows (infection-centered vs ICU-admission-centered) and infection detection methods (antibiotic-culture matching vs APACHE-based diagnosis). Dependencies among several MIMIC-III repositories suggested propagation of implementation decisions across studies.

**Discussion:** Clinically validated sepsis definitions yield substantially different detection rates across studies using identical datasets, indicating heterogeneity in computational implementation.

**Conclusion:** To improve reproducibility in sepsis research and the robustness of sepsis prediction models, we recommend standardized reporting of sepsis case detection methodology and the publication of version-controlled source code.

## Background and Significance

Sepsis is a major clinical and economic burden globally, accounting for 11 million deaths in 2017 and representing 19 % of global mortality [1, 2]. In intensive care settings, sepsis affects 29.5 % of patients, and mortality rates exceed 35 % [3, 4], resulting in substantial healthcare costs [5, 6]. Clinical guidelines emphasize systematic screening for early detection [7], and the introduction of the Sepsis-3 definition in 2016 marked an important step toward standardization [8]. This definition operationalizes sepsis as suspected infection accompanied by acute organ dysfunction. Organ dysfunction is quantified with the SOFA (Sequential Organ Failure Assessment) score, and an increase of at least 2 SOFA points indicates sepsis when a concomitant infection is apparent. Nevertheless, applying this definition to identical datasets yields substantially different sepsis detection rates across studies [9].

This variability likely reflects the complexity inherent in algorithmic implementations of automated sepsis case detection. The SOFA score requires the integration of six organ system assessments (respiratory, cardiovascular, hepatic, coagulation, renal, and neurological), each dependent on clinical parameters that may be recorded under multiple labels, measured at irregular intervals, and subject to data quality issues including missing values and documentation inconsistencies. Automated detection of infections compounds this complexity by requiring either ICD-based diagnoses or temporal matching of antibiotic administration with microbiology culture sampling [10].

The scale of methodological heterogeneity is evident when examining studies applying the current sepsis definition to the Medical Information Mart for Intensive Care III (MIMIC-III [11]). Despite utilizing identical source data and nominally following the same definition, reported detection rates range from 11.3 % to 49.1 % [12]. Lambden et al. [9] highlighted challenges in sepsis operationalization, yet a systematic characterization of the specific implementation decisions contributing to this variability is lacking.

Two methodological gaps contribute to these challenges. First, publications frequently omit sepsis case definition details such as parameter identification strategies, temporal aggregation methods, missing value imputation approaches, SOFA calculation variants and suspicion of infection details, thereby making exact reproduction difficult. Second, without consensus on best practices for retrospective sepsis case operationalization, researchers make divergent methodological choices across multiple dimensions of data preprocessing and label calculation.

This study addresses these gaps through a systematic, multi-level investigation of sepsis case detection methodologies in MIMIC-III [11] and eICU-CRD [13] databases. We combined a PRISMA-guided systematic literature review with detailed source code analysis to answer three research questions:

1. What is the range of sepsis detection rates reported across studies applying Sepsis-3 to MIMIC-III/eICU-CRD?
2. Which methodological decisions in data preprocessing and label calculation can be identified from publications and source code?
3. What patterns of methodological heterogeneity emerge across implementations, and to what extent are implementation decisions documented?

To structure this analysis, we decomposed sepsis detection decisions into six methodological domains (D1-D6): parameter coverage (D1), temporal windows (D2), aggregation strategies (D3), missing value handling (D4), SOFA calculation variants (D5), and infection detection methods (D6).

## Materials and Methods

We first identified publicly available intensive care databases (MIMIC-III [11], MIMIC-IV [14], eICU-CRD [13], AmsterdamUMCdb [15]). For each database, we conducted literature searches in PubMed and Web of Science to identify sepsis related publications. Separately, we compiled a corpus of publications related to the current sepsis definition. We then computed the intersection of each database-specific set with the sepsis corpus to quantify the volume of sepsis detection studies per database. MIMIC-III yielded the largest intersection and was therefore selected for detailed analysis. We also included eICU-CRD as the second dataset for three reasons: (1) sufficient publication volume to enable systematic analysis (110 sepsis publications), (2) healthcare system comparability with MIMIC-III (both US-based), and (3) institutional independence from MIMIC-III, ensuring no patient data overlap. MIMIC-IV was excluded despite having the second-largest corpus (264 publications) because it originates from the same institution as MIMIC-III with overlapping patient populations (2008–2012), precluding independent validation. AmsterdamUMCdb was excluded due to insufficient publication volume (17 sepsis publications).

The Medical Information Mart for Intensive Care III (MIMIC-III, version 1.4) contains de-identified electronic health record data from 46,520 patients admitted to intensive care units at Beth Israel Deaconess Medical Center between 2001 and 2012 [11]. MIMIC-III integrates data from two distinct clinical information systems: *CareVue* (primarily 2001–2008) and *MetaVision* (2008–2012), which differ in parameter naming conventions, documentation granularity, and data completeness [16]. The eICU Collaborative Research Database (eICU-CRD, version 2.0) contains de-identified electronic health record data from 200,859 ICU stays corresponding to 139,367 unique patients admitted to 208 hospitals in the United States during 2014–2015 [13].

We searched PubMed and Web of Science from January 1, 2016 through December 31, 2024, covering the period since the Sepsis-3 definition was published. Search terms for MIMIC-III and Sepsis-3 comprised spelling variations as well as citation-based searches of the original Sepsis-3 definition [8] or MIMIC-III [11] publications. Complete search strategies with counts of resulting publications are provided in Supplement S1. Studies were included if they: (1) were published in English between January 2016 and December 2024, (2) were available as open access full text, (3) used the MIMIC-III or eICU dataset with explicit SOFA-based sepsis detection, and (4) provided label calculation details, and (5) reported a traceable sepsis detection rate. A study provided label calculation details if, beyond applying the Sepsis-3 definition, it described at least one concrete step in the label calculation (e.g., parameter aggregation, the observation window, imputation, or outlier handling), the infection-identification method, or a link to source code. A rate was traceable when a septic count and a cohort size could be obtained in a consistent unit, whether reported or reconstructed from the text, appendix, or a flow diagram, even without a documented infection detection method. Examples are given in Supplement S1. We excluded review articles, conference abstracts without full papers, studies mentioning but not using MIMIC-III/eICU-CRD or the current sepsis definition (Sepsis-3), and those without any traceable sepsis cohort derivation. The selection process followed PRISMA guidelines through three phases: initial title/abstract screening, full-text assessment for eligibility, and final inclusion. Abstract screening was performed by three independent reviewers, with discrepancies resolved through discussion. To detect suspected infection from clinical data retrospectively, it was investigated whether implementations adhered to ICD or other methods, like the antibiotic-culture temporal matching method proposed by Seymour et al. [10].

Where a study narrowed to a clinical subgroup (a specific disease, organ dysfunction, or comparable clinical condition; e.g., sepsis-associated acute kidney injury), we included it only if it reported a general adult sepsis cohort before the narrowing, from which a comparable baseline detection rate could be recovered. If the cohort was restricted to the subgroup before sepsis identification, leaving only a subgroup rather than a general adult sepsis rate, we excluded the study (hidden conditional filtering). This differs from the data-quality or processing filters applied by some included studies (e.g., missingness thresholds, outlier removal, or a minimum observation window), which narrow the general adult cohort without redefining the clinical population. Studies were excluded for incomparable sepsis labels if they did not provide a single sepsis cohort that was directly comparable with those from the other included studies. This included studies reporting multiple non-comparable cohort estimates (e.g., using different onset definitions) without an objective basis for selecting one, or studies based on systematically different source populations or data subsets that were not comparable with the reference cohorts.

We developed a data extraction protocol covering six methodological domains (D1–D6) that span the complete process from raw clinical data to final sepsis case detection. Parameter coverage (D1) encompasses clinical parameter selection and name variants, mechanical ventilation parameters, antibiotic names/routes for infection detection, and dual information system handling (*CareVue*/*MetaVision*). Temporal resolution (D2) covers SOFA calculation scope, sliding window size/offset, and temporal reference points (ICU admission vs. infection onset). Parameter aggregation (D3) includes strategies for multiple measurements within windows (worst-case vs. central tendency), cumulative parameters (rolling sums for urine output), and vasopressor aggregation. Missing data handling (D4) encompasses imputation methodologies, fallback strategies, formula-based calculations, and patient-specific vs. population-based approaches. SOFA calculation variants (D5) include baseline assumptions and component-specific modifications. Infection detection methods (D6) cover primary approaches (ICD-9 codes, antibiotic-culture matching [10], hybrid methods) and antibiotic list comprehensiveness.

In addition, we documented cohort selection criteria including age restrictions, ICU stay duration requirements, exclusion thresholds for patients with excessive data missingness, multiple admission handling policies, and early sepsis exclusion strategies. For each study, we extracted reported cohort sizes and sepsis detection rates (as patients, admissions, or ICU stays), and technical implementation details. Among the included studies, in order to facilitate interpretation of detection rate variations, we stratified them based on cohort comparability: Set A Studies reported adult sepsis detection rates on standard reference populations, without any other additional filters beyond age restrictions; Set B Studies applied additional filters other than clinical-subgroup restrictions (e.g., missingness thresholds, outlier removal, minimum observation duration).

We additionally categorized each included study from its abstract as an observational (O), prediction-model (P), treatment-effect (T), or methods (M) study, following the reporting and methodological frameworks that anchor these categories: STROBE and PROGRESS for observational studies [17, 18], TRIPOD and TRIPOD+AI for prediction models [19, 20], target-trial emulation for treatment-effect studies [21], and frameworks for methodological and neutral-comparison research for methods studies [22, 23]. As study design was uniformly retrospective, this categorization served as a study-level covariate in the detection-rate analysis. The assigned category is given per study in Tables 1 and 2.

**Table 1.** Sepsis detection rates across 44 MIMIC-III/Sepsis-3 studies. Set A (No. 1–24): standard reference populations without additional data quality filters; Set B (No. 25–44): additional filtering applied. Entity indicates the cohort source (patients, admissions, ICU stays, or the MetaVision subset); # all = total cohort size; # septic = sepsis cases detected. Values in brackets were inferred from methodology or cited references. Abbreviations: ATB+Lab = antibiotics plus microbiology cultures [10]; ICD9 = ICD-9 diagnosis codes; Angus = Angus criteria; NR = not reported. Reference sizes: Patients *n*=46,520; Admissions *n*=58,976; ICU Stays *n*=61,532. Code: ✓ = available (^1^original, ^2^derived, ^3^MIMIC repository [28]). NR for infection denotes an unreported infection detection method and does not imply an untraceable detection rate. The Type column indicates each study’s classification as observational (O), methods (M), prediction model (P), or treatment effect (T), as defined in the Methods.

| No. | Author | Type | Code | Version | Entity | Min. Age | Infection | # all | # septic | Rate |
| --- | --- | --- | --- | --- | --- | --- | --- | --- | --- | --- |
| *Set A – Standard reference populations* |  |  |  |  |  |  |  |  |  |  |
| 1 | Wang et al. [29] | O |  | 1.4 | Patients | 16 | ICD9+Lab | [38,641] | 23,956 | 62.00 % |
| 2 | Parbhoo et al. [30] | M | ✓ | 1.4 | [Patients] | 15 | NR | [38,605] | 18,200 | 47.14 % |
| 3 | Peng et al. [31] | O |  | 1.4 | Patients | – | SIC score | [46,520] | 19,613 | 42.16 % |
| 4 | Chen et al. [32] | O |  | 1.4 | Patients | – | ICD9 | 46,520 | 18,592 | 39.97 % |
| 5 | Yao et al. [33] | P |  | [1.4] | Patients | 16 | ICD9 | 38,641 | 15,032 | 38.90 % |
| 6 | Chen et al. II [34] | O |  | 1.4 | Patients | 15 | NR | 38,605 | 14,861 | 38.50 % |
| 7 | Drudi et al. [35] | T |  | [1.4] | [Admissions] | 16 | ATB+Lab | 53,423 | 20,496 | 38.37 % |
| 8 | Huang et al. [36] | O |  | 1.4 | [Admissions] | 16 | NR | 53,423 | 20,438 | 38.26 % |
| 9 | Chen et al. III [37] | O |  | [1.4] | [Admissions] | 16 | ICD9 | 53,423 | 18,592 | 34.80 % |
| 10 | Choi et al. [38] | T |  | 1.4 | [ICU Stays] | – | NR | 61,532 | 20,927 | 34.01 % |
| 11 | Chen et al. IV [39] | P |  | 1.4 | [ICU Stays] | 14 | ATB+Lab | 52,961 | 13,828 | 26.11 % |
| 12 | Wang et al. [40] | P |  | [1.4] | [Patients] | – | ATB+Lab | [46,520] | 11,791 | 25.35 % |
| 13 | Shahn et al. [41] | T |  | [1.4] | MetaVision | 18 | ATB+Lab | 23,606 | 5,784 | 24.50 % |
| 14 | Zhang et al. [42] | P |  | [1.4] | MetaVision | – | NR | 23,630 | 5,443 | 23.03 % |
| 15 | Hou et al. [43] | P |  | 1.4 | Patients | – | ICD9 | 46,520 | 10,704 | 23.01 % |
| 16 | Ren et al. [44] | P |  | 1.4 | [Patients] | – | ICD9 | [46,520] | 7,851 | 16.88 % |
| 17 | Rosnati et al. [45] | P | ✓ | [1.4] | Admissions | 15 | ATB+Lab | NR | 7,936 | [14.86] % |
| 18 | Zhou et al. [46] | P |  | [1.4] | [Admissions] | 18 | ATB+Lab | [52,862] | 7,230 | 13.68 % |
| 19 | Peng et al. [47] | P |  | [1.4] | [Admissions] | 18 | ICD9 | [52,862] | 5,784 | 10.94 % |
| 20 | Yan et al. [48] | O |  | [1.4] | Admissions | 16 | NR | 52,963 | 4,727 | 8.93 % |
| 21 | Apalak & Kiasaleh [49] | P |  | 1.4 | MetaVision | 15 | ATB+Lab | 19,515 | 1,725 | 8.84 % |
| 22 | Lan et al. [50] | O |  | 1.4 | Patients | 16 | ICD9 | [38,641] | 1,891 | 4.89 % |
| 23 | Wang et al. II [51] | O |  | 1.4 | [ICU Stays] | – | NR | 61,532 | 2,733 | 4.44 % |
| 24 | Sakib et al. [52] | O |  | 1.4 | [Admissions] | – | ATB+Lab | 58,976 | 1,994 | 3.38 % |
| *Set B – Additional filtering applied* |  |  |  |  |  |  |  |  |  |  |
| 25 | Lin et al. [53] | O |  | 1.3 | Patients | 18 | NR | 18,894 | 12,321 | 65.21 % |
| 26 | Zhong et al. [54] | O |  | [1.4] | [Patients] | – | [ICD9] | 38,597 | 20,318 | 52.64 % |
| 27 | Wang et al. III [55] | O |  | [1.4] | [ICU Stays] | 16 | ICD-9 (Angus) | 31,951 | 16,131 | 50.49 % |
| 28 | Johnson et al. [56] | M | ✓ | 1.4 | MetaVision | 18 | ATB+Lab | 11,791 | 5,784 | 49.05 % |
| 29 | Wang et al. [57] | O |  | 1.4 | MetaVision | 16 | ATB+Lab | 13,783 | 5,783 | 41.96 % |
| 30 | Liu et al. [58] | P |  | 1.4 | [Patients] | 18 | ATB+Lab | 38,418 | 15,111 | 39.33 % |
| 31 | Ghalati et al. [59] | M |  | 1.4 | Admissions | – | ATB+Lab | 58,976 | 22,547 | 38.23 % |
| 32 | Rhodes et al. [60] | M | ✓<sup>2</sup> | [1.4] | [ICU Stays] | [18] | [ATB+Lab] | NR | 20,954 | [34.05] % |
| 33 | Rhodes et al. [61] | M | ✓<sup>2</sup> | 1.4 | [ICU Stays] | [18] | [ATB+Lab] | NR | 20,952 | [34.05] % |
| 34 | Liu et al. [62] | T | ✓ | [1.4] | [Patients] | 18 | ATB+Lab | 38,645 | 10,840 | 28.05 % |
| 35 | Komorowski et al. [63] | T | ✓<sup>1</sup> | [1.4] | [ICU Stays] | 18 | ATB+Lab | NR | 17,083 | [27.76] % |
| 36 | Chen et al. [64] | O | ✓ | 1.4 | [Patients] | 18 | NR | NR | 10,305 | [26.67] % |
| 37 | Moor et al. [65] | P | ✓ | 1.4 | Admissions | 15 | ATB+Lab | 36,591 | 9,541 | 26.07 % |
| 38 | He & Qiu [66] | P | ✓<sup>3</sup> | 1.4 | [Admissions] | 18 | ATB+Lab | NR | 7,924 | [14.67] % |
| 39 | Wei et al. [67] | O |  | 1.4 | [Patients] | 18 | [ICD9] | 38,645 | 5,325 | 13.78 % |
| 40 | Peng et al. [68] | O |  | 1.4 | Admissions | 18 | NR | 43,129 | 5,784 | 13.41 % |
| 41 | Desautels et al. [16] | P |  | 1.3 | MetaVision | 15 | ATB+Lab | 22,853 | 2,577 | 11.28 % |

Table 1 continued from previous page
| No. | Author | Type | Code | Version | Entity | Min. Age | Infection | # all | # septic | Rate |
| --- | --- | --- | --- | --- | --- | --- | --- | --- | --- | --- |
| 42 | Li et al. [69] | O |  | 1.3 | Patients | 18 | NR | 46,476 | 5,121 | 11.02 % |
| 43 | Solís-García et al. [70] | M |  | 1.4 | MetaVision | 15 | ATB+Lab | 19,073 | 1,797 | 9.42 % |
| 44 | Barton et al. [71] | P |  | 1.3 | [ICU Stays] | 18 | ATB+Lab | 21,507 | 1,024 | 4.76 % |

**Table 2.** Sepsis detection rates across 20 eICU-CRD/sepsis studies. Entity indicates the cohort source; # all = total cohort size; # septic = sepsis cases detected. Values in brackets were inferred from methodology or cited references. Reference sizes: Patients *n*=139,367; ICU Stays *n*=200,859. Abbreviations: ATB+Lab = antibiotics plus microbiology cultures [10]; ICD9 = ICD-9 diagnosis codes; NR = not reported; doc = documented; sus = suspected; con = confirmed; kno = known; inf = infection. Code: ✓ = available (^1^original, ^2^derived, ^3^MIT-LCP repository). ^4^No further infection information is provided. ^5^Total cohort size including patients under the min. age. NR for infection denotes an unreported infection detection method and does not imply an untraceable detection rate. The Type column indicates each study’s classification as observational (O), methods (M), prediction model (P), or treatment effect (T), as defined in the Methods.

| No. | Author | Type | Code | Version | Entity | Min. Age | Infection | # all | # septic | Rate |
| --- | --- | --- | --- | --- | --- | --- | --- | --- | --- | --- |
| 1 | Fang et al. [72] | O |  | [2.0] | [ICU Stays] | [18] | – | [200,859]<sup>5</sup> | 96,127 | 47.86 % |
| 2 | Zhang et al. [73] | P |  | 2.0 | Patients | 18 | doc./sus. inf.<sup>4</sup> | [139,367]<sup>5</sup> | 48,694 | 34.94 % |
| 3 | Zhang et al. [74] | O |  | 2.0 | Patients | 18 | con./sus. inf.<sup>4</sup> | [139,367]<sup>5</sup> | 48,694 | 34.94 % |
| 4 | Zhang et al. [75] | O |  | 2.0 | Patients | 18 | con. inf.<sup>4</sup> | [139,367]<sup>5</sup> | 48,694 | 34.94 % |
| 5 | Zhang et al. [76] | P | ✓<sup>3</sup> | 2.0 | Patients | – | – | [139,367] | 43,727 | 31.38 % |
| 6 | Yang et al. [77] | O |  | 2.0 | [ICU Stays] | 18 | – | 200,859<sup>5</sup> | 55,848 | 27.80 % |
| 7 | Zhang et al. [78] | O |  | – | Patients | – | inf.<sup>4</sup> | [139,367] | 36,302 | 26.05 % |
| 8 | Wang et al. [79] | O | ✓<sup>3</sup> | – | Patients | – | inf.<sup>4</sup> | [139,367] | 36,046 | 25.86 % |
| 9 | Li et al. [80] | O |  | 2.0 | Patients | – | ATB+Lab | [139,367] | 31,737 | 22.77 % |
| 10 | Zhao et al. [81] | P |  | – | ICU Stays | – | ATB+Lab | 200,859 | 41,667 | 20.74 % |
| 11 | Lin et al. [82] | O |  | [2.0] | Patients | – | con. inf.<sup>4</sup> | [139,367] | 23,136 | 16.60 % |
| 12 | Lin et al. [83] | O |  | [2.0] | Patients | – | ICD-9 | [139,367] | 23,136 | 16.60 % |
| 13 | Shen et al. [84] | P |  | 2.0 | Patients | – | – | [139,367] | 23,136 | 16.60 % |
| 14 | Chang et al. [85] | O |  | – | Patients | – | ICD-9 | [139,367] | 23,136 | 16.60 % |
| 15 | Yang et al. [86] | P | ✓<sup>1</sup> | – | Patients | – | kno./sus. inf.<sup>4</sup> | [139,367] | 22,864 | 16.41 % |
| 16 | Kasugai et al. [87] | O |  | – | [ICU Stays] | – | doc./sus. inf.<sup>4</sup> | 200,859 | 30,114 | 14.99 % |
| 17 | Wang et al. [88] | O | ✓<sup>2</sup> | 2.0 | ICU Stays | – | – | 200,859 | 28,667 | 14.27 % |
| 18 | Li et al. [89] | O |  | 2.0 | Patients | – | – | [139,367] | 19,430 | 13.94 % |
| 19 | Sheng et al. [90] | O |  | 2.0 | [ICU Stays] | [18] | – | 200,859<sup>5</sup> | 26,360 | 13.12 % |
| 20 | Takkavatakarn et al. [91] | O | ✓<sup>3</sup> | [2.0] | ICU Stays | [18] | ATB+Lab | [200,859]<sup>5</sup> | 19,575 | 9.75 % |

To quantify detection-rate variability, we fitted a one-stage generalized linear mixed model (GLMM) to the 58 studies with both a septic count and a cohort size. This binomial–normal random-effects model treats each study’s septic count as binomial given its cohort size, with a normal study-level random intercept on the logit scale. We preferred it to the transformation-based two-stage DerSimonian–Laird method [24], whose moment estimator underestimates the between-study variance (*τ*^2^) when heterogeneity is high—as it is here—whereas the GLMM estimates *τ*^2^ directly from the exact binomial likelihood [25]; this DerSimonian–Laird analysis is reported as a sensitivity check (Supplement S1, Section 6). From the GLMM we report *τ*^2^ (with a profile-likelihood CI) and the 95 % prediction interval, overall and within each database. Because each cohort is very large, Cochran’s *Q* and *I*^2^ (with 95 % CI [26]) are near their maxima by construction, so inference was based on *τ*^2^ and the prediction interval. We further compared the per-study detection rates across the database, unit of analysis, cohort set, study type, infection detection method and code availability using the Wilcoxon rank-sum test for two-level characteristics and the Kruskal–Wallis test otherwise. A random-effects meta-regression, using the same GLMM, included the first three as pre-specified covariates and added each of the remaining three individually. Analyses used the R packages lme4 and metafor and the Python packages NumPy and SciPy; the GLMM was fitted in lme4 and cross-checked against metafor, and both gave identical estimates.

We systematically identified source code through two channels: code repository links provided in publications identified through our systematic search, and supplementary material of included publications. In addition, dependency tracing revealed upstream repositories containing complete Sepsis-3 implementations not captured by the literature search. We selected code repositories meeting three additional criteria: (1) peer-reviewed publication association with studies identified in our systematic literature search, (2) explicit MIMIC-III/eICU-CRD and SOFA-based sepsis detection implementation verified through source code inspection, and (3) sufficient code completeness for methodology extraction. When re-implementations were identified, we traced code sources back to original implementations. For each identified code repository, we applied the same six-dimensional methodological framework (D1 to D6) to extract implementation details directly from source code.

All analyzed repositories were publicly accessible. Complete documentation is provided in Supplement S2, with a summary in the Results Section. Claude Code [27] (Anthropic Claude Opus 4.5, accessed January 2026, and Opus 4.8, accessed June 2026) was used to assist with (1) structuring and organizing findings from the source code analysis, (2) generating code for data management and figure preparation, and (3) language editing and refinement of the manuscript and supplement text. All AI-generated outputs were manually verified and validated by the authors.

## Results

### Systematic Literature Search

For MIMIC-III, the literature search identified 262 records from Web of Science and 115 from PubMed at the intersection of MIMIC-III and sepsis related publications, supplemented by 8 records from reference screening of seminal articles. After duplicate removal, 286 unique papers remained for screening. For eICU-CRD, a parallel search yielded 110 unique papers after duplicate removal. In total, 396 papers were screened across both databases (M: 286; E: 110), including 32 articles appearing in both searches (Figure 1).

**Figure 1.**
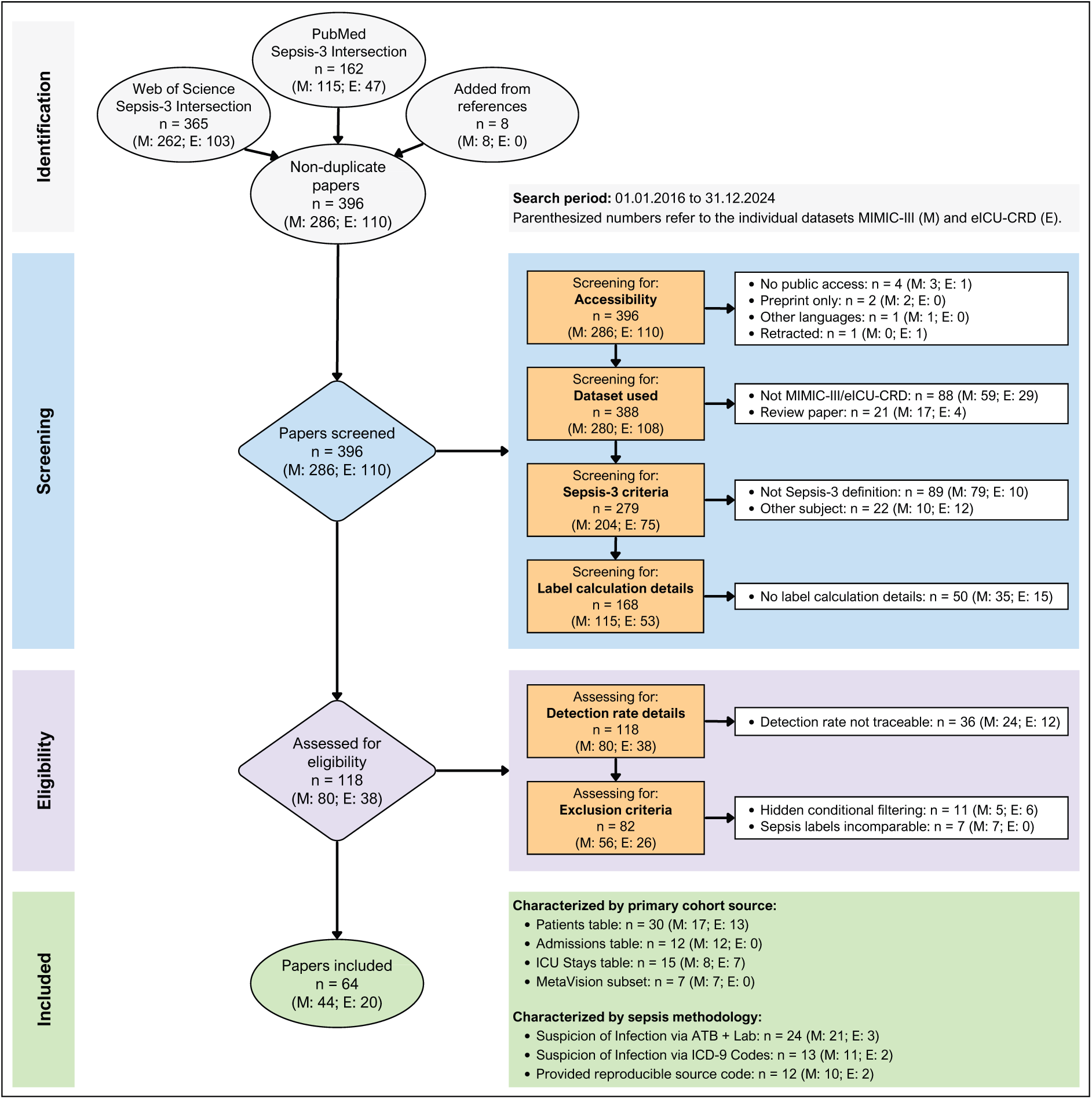
PRISMA flow diagram of the study selection process for MIMIC-III (M) and eICU-CRD (E) datasets. The diagram illustrates the phases of identification, screening, eligibility assessment, and final inclusion. After duplicate removal, 396 papers were screened (M: 286; E: 110). Articles were sequentially excluded based on accessibility, dataset used, adherence to current sepsis criteria, and provision of label calculation details, yielding 118 articles for eligibility assessment (M: 80; E: 38). Following evaluation of detection rate traceability and exclusion criteria comparability, 64 studies were included for detailed analysis (M: 44; E: 20).

During the screening phase, articles were sequentially excluded based on four criteria: accessibility (records without open access, preprint-only status, or non-English language), dataset used (non-MIMIC-III/eICU-CRD studies and review papers), adherence to current sepsis criteria (studies not implementing SOFA-based sepsis detection or with unrelated subjects), and provision of label calculation details. This yielded 118 articles for eligibility assessment (MIMIC-III: 80; eICU-CRD: 38). In the eligibility phase, articles were further evaluated for traceability of reported sepsis detection rates and comparability of exclusion criteria (excluding studies with hidden conditional filtering or incomparable sepsis labels). The final corpus comprised 64 studies for detailed analysis (MIMIC-III: 44; eICU-CRD: 20).

### Sepsis Detection Rate Variability Across Studies

Table 1 presents detection rates, cohort sizes, and key methodological characteristics for all 44 MIMIC-III studies, stratified into Set A (No. 1–24, n=24) and Set B (No. 25–44, n=20).

Across all 44 MIMIC-III studies, reported sepsis detection rates ranged from 3.4 % to 65.2 % (median: 26.4 %, IQR: 13.6 %–38.6 %). Set A studies reported rates from 3.4 % to 62.0 % (median: 24.9 %, IQR: 13.0 %–38.4 %), while Set B studies showed comparable variability from 4.8 % to 65.2 % (median: 27.9 %, IQR: 13.7 %–40.0 %).

To assess generalizability beyond MIMIC-III, we identified 20 additional publications implementing the current sepsis definition on the eICU-CRD (Table 2), which aggregates data from 208 US hospitals. All 20 eICU-CRD studies reported detection rates based on the general adult sepsis cohort prior to applying study-specific exclusion criteria, corresponding to Set A.

The 20 eICU-CRD studies showed rates ranging from 9.8 % to 47.9 % (median: 18.7 %, IQR: 16.1 %–28.7 %). Comparing the two databases, MIMIC-III studies showed a numerically higher median detection rate (26.4 % vs. 18.7 %; not statistically significant, see below) and wider variability (range: 61.8 vs. 38.1 percentage points; IQR: 25.0 vs. 12.6 percentage points).

Variability persisted even among studies analyzing identical source populations within each database. In MIMIC-III, five studies deriving cohorts from the patients table without age restrictions (n=46,520) reported detection rates ranging from 16.9 % to 42.2 %. Similarly, in eICU-CRD, ten studies analyzing the full patients table (n=139,367) without age restrictions reported detection rates from 13.9 % to 31.4 %. Since these studies operated on identical source data, this within-population variability demonstrates that implementation decisions, rather than patient selection, contribute to detection rate differences. Detection rates showed no trend toward convergence over time in either database (Figure 2; MIMIC-III: slope = −0.4 %/year, p = 0.75; eICU-CRD: slope = +1.7 %/year, p = 0.25).

**Figure 2.**
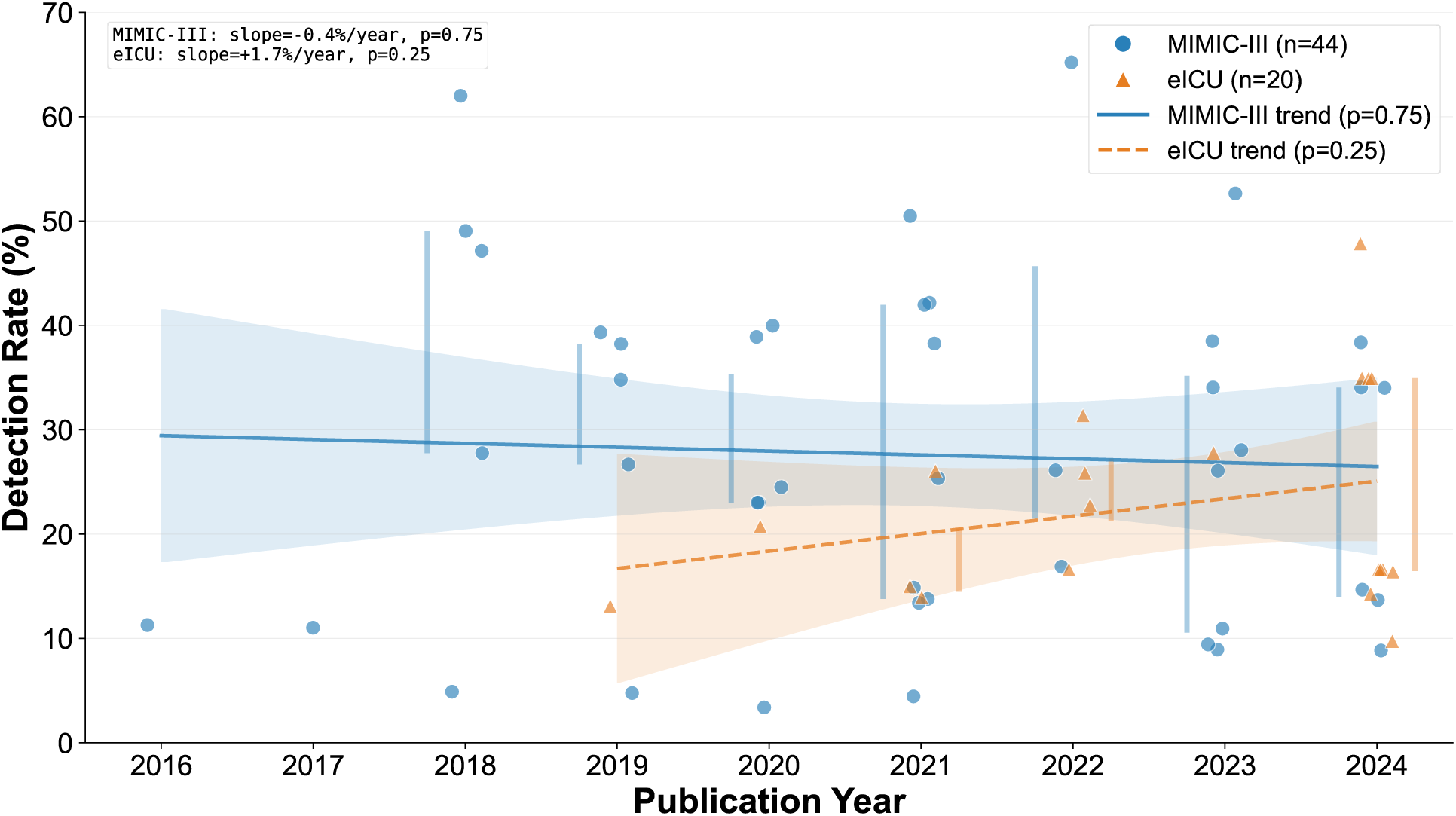
Detection rates by publication year across MIMIC-III (n=44, blue circles) and eICU (n=20, orange triangles) studies (2016–2024). Each point represents one study; points are jittered horizontally for visibility. Lines show linear regression trends with 95 % confidence bands (shaded areas). Vertical bars indicate interquartile ranges for years with ≥ 3 studies.

Figure 3 shows the study-level detection rates stratified by the database and study characteristics. The distributions overlapped substantially within every subgroup, and no characteristic separated the rates (Wilcoxon rank-sum or Kruskal–Wallis tests, *p* 0.17–0.70; Supplement S1, Section 6).

**Figure 3.**
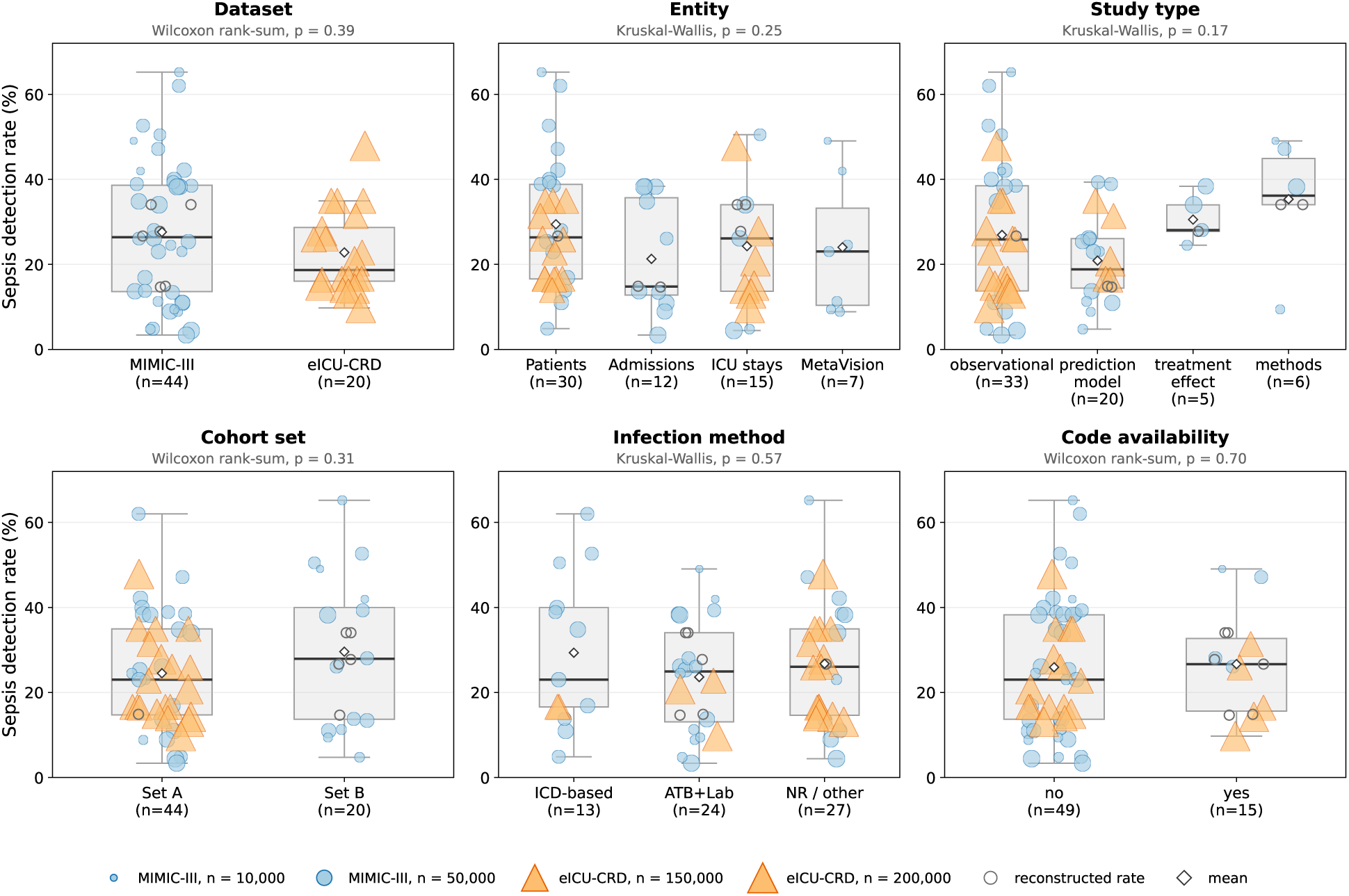
Study-level sepsis detection rates by study characteristic. Each panel stratifies the rates by one characteristic: database, entity (unit of analysis), study type, cohort set, infection detection method and code availability. Boxplots show the median (line), mean (diamond) and interquartile range; overlaid markers are individual studies, with area proportional to cohort size and shape denoting the database (circles, MIMIC-III; triangles, eICU-CRD). Open markers denote studies whose detection rate was reconstructed from an assumed denominator. Above each panel is the nonparametric test across levels (Wilcoxon rank-sum for two-level characteristics, Kruskal–Wallis otherwise); the boxplots and tests use all 64 studies.

Of the 64 studies, the six whose detection rate was reconstructed from an assumed denominator were excluded from the meta-analysis but shown in Figure 3 as open markers, leaving 58. Within each database the detection rates were highly heterogeneous (from the GLMM: MIMIC-III *τ*^2^ = 1.05, eICU-CRD *τ*^2^ = 0.29 on the logit scale; Table 3); because the cohorts are very large, sampling error was negligible and essentially all of this variation lay between studies. The two databases did not differ significantly (Wilcoxon rank-sum *p* = 0.39). The meta-regression explained little of the overall between-study variance (*τ*^2^ from 0.79 to 0.72), and no characteristic was significantly associated with the rate; the largest difference was for the unit of analysis, with admission- and ICU-stay cohorts showing lower rates than patient cohorts (odds ratios ≈ 0.6, all *p* ≥ 0.08), a mechanical effect of the changing denominator. Findings were robust across sensitivity analyses (Supplement S1, Section 6).

**Table 3.**
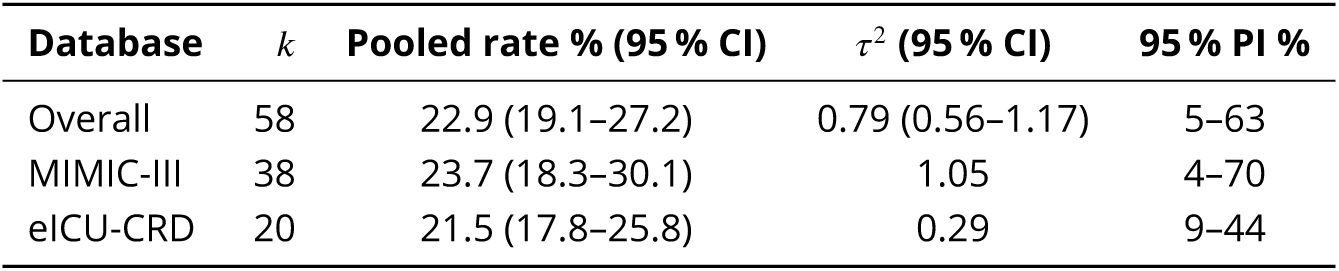
Between-study heterogeneity of sepsis detection rates from the one-stage binomial–normal GLMM. *k*, number of studies; pooled rate with 95 % CI; *τ*^2^, between-study variance on the logit scale (95 % profile-likelihood CI for the overall estimate); 95 % PI, prediction interval for a new study. *I*^2^ > 99.9 % in all rows (saturated by the very large cohorts), so inference is based on *τ*^2^ and the prediction interval. Robustness checks, the DerSimonian–Laird sensitivity analysis, and the meta-regression are in Supplement S1, Section 6.

Among MIMIC-III studies, ten (22.7 %) provided links to publicly accessible source code, with code availability notably higher among Set B studies (8/20, 40.0 %) compared to Set A (2/24, 8.3 %). Two eICU-CRD studies (10.0 %) provided source code, three additional studies were linked to an upstream MIT-LCP repository through dependency tracing (Table 2).

### Methodological Heterogeneity Across Publications

Analysis of preprocessing methodology across 44 MIMIC-III/sepsis studies revealed documentation gaps and methodological heterogeneity across all five domains examined (D2–D6). Parameter coverage (D1) could not be reliably assessed from publication text alone and is addressed through source code analysis in the next section. Documentation rates, defined as explicit reporting of methods applicable to sepsis label calculation, were uniformly low: SOFA calculation parameters (34.1 %, n=15), infection detection methods (31.8 %, n=14), temporal windows (15.9 %, n=7), aggregation methods (11.4 %, n=5), and missing data handling (6.8 %, n=3). A significant proportion of studies described preprocessing steps where applicability to label calculation versus prediction model development remained ambiguous (D2: 15.9 %, D3: 20.5 %, D4: 47.7 %), complicating reproducibility assessment. Regarding temporal resolution (D2), among the 7 studies with explicit temporal documentation for label calculation, aggregation window sizes ranged from 1 hour to 24 hours. For missing data handling (D4), only 3 studies explicitly documented methods for label calculation, all combining forward-fill with default-to-normal assumptions for missing SOFA components – a practice that may reduce SOFA scores and thus underestimate sepsis detection rates. Regarding SOFA calculation (D5), among the 15 studies with explicit documentation for label calculation, 12 stated the ≥ 2 threshold required by the current sepsis definition. However, baseline SOFA determination, critical for detecting the 2-point increase rather than absolute score, was documented in only 2 studies. For infection detection methods (D6), among the 14 studies with explicit documentation, the antibiotic-culture temporal matching criteria [10] predominated (n=11), with ICD-9 diagnostic codes used in 2 studies.

Figure 4 illustrates the relationship between documented preprocessing methods and sepsis detection rates across all domains. Substantial variability exists within each methodological category, suggesting that documented preprocessing choices alone do not explain the observed detection rate heterogeneity.

**Figure 4.**
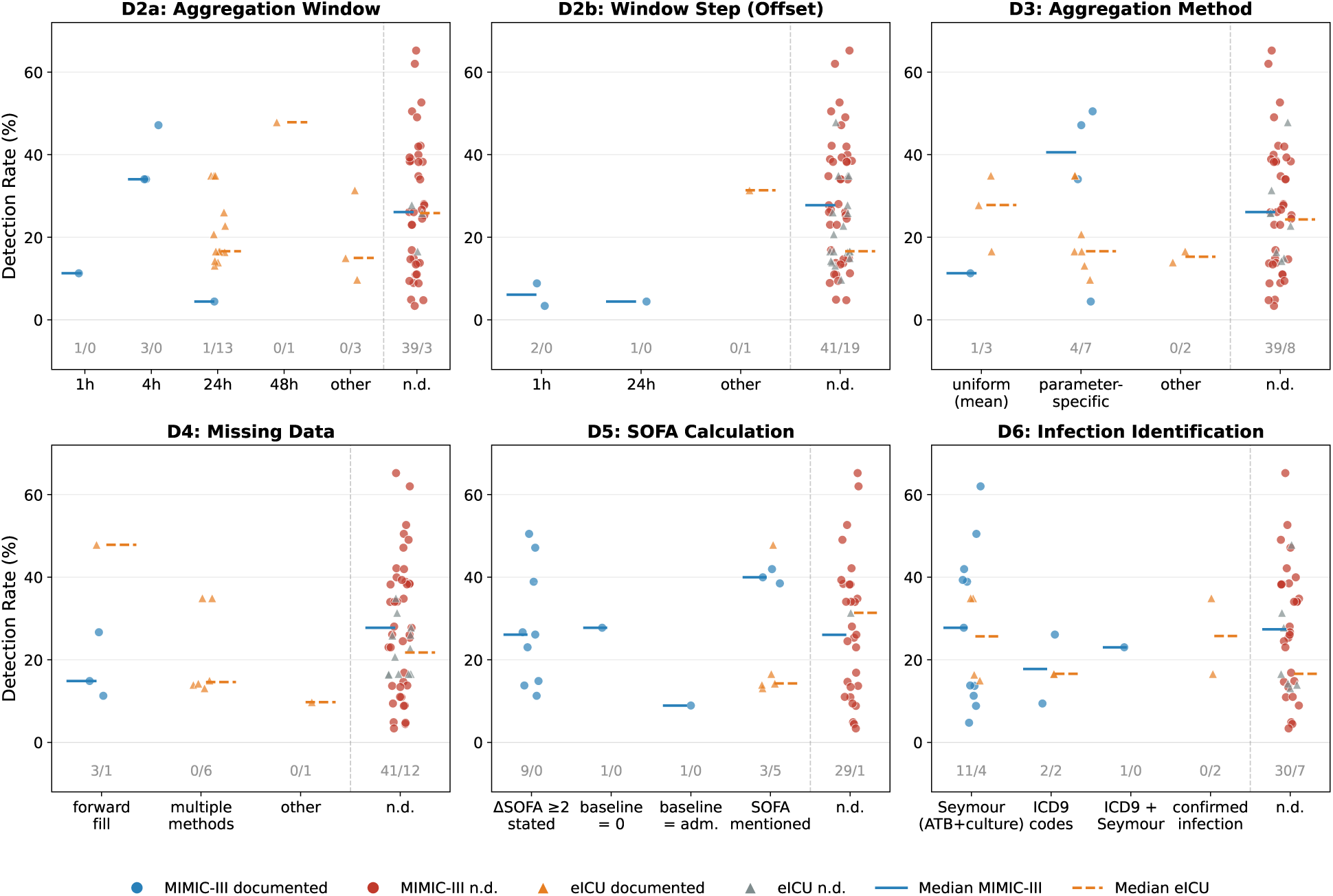
Detection rate variability across preprocessing methodology in MIMIC-III (n=44) and eICU-CRD (n=20) studies. Panels show temporal windows (D2a/b), aggregation methods (D3), missing data handling (D4), SOFA calculation (D5), and infection detection methods (D6). Blue circles: MIMIC-III documented; red circles: MIMIC-III not documented (n.d.); orange triangles: eICU-CRD documented; gray triangles: eICU-CRD n.d. Solid lines: MIMIC-III medians; dashed lines: eICU-CRD medians. Sample sizes shown as MIMIC-III/eICU-CRD above each category.

Documentation rates in eICU-CRD studies (n=20) were considerably higher: SOFA calculation (95.0 %, n=19), infection detection methods (65.0 %, n=13), temporal windows (85.0 %, n=17), aggregation methods (60.0 %, n=12), and missing data handling (40.0 %, n=8). Despite higher documentation, detection rate variability persisted (range: 38.1 percentage points), confirming that methodological heterogeneity rather than documentation alone drives outcome differences. Across both databases (n=64), combined documentation rates were: SOFA calculation (53.1 %), infection detection methods (42.2 %), temporal windows (37.5 %), aggregation methods (26.6 %), and missing data handling (17.2 %). Parameter coverage (D1) was assessed exclusively through source code analysis.

### Source Code Analysis

Of the 64 shortlisted articles, 12 provided references to openly accessible source code, though several referenced identical repositories. After deduplication and screening for complete Sepsis-3 label calculation implementations, six repositories remained for detailed analysis: Johnson et al. [56] (sepsis3-mimic; 18 files, 110 decisions), Komorowski et al. [63] (AI_Clinician; 3 files, 36 decisions), Johnson et al. [28] (mimic-code; 34 files, 50 decisions), and Rosnati et al. [45] (MIMIC-III-sepsis-3-labels; 25 files, 68 decisions) for MIMIC-III; Chen et al. [92] (16 files, 34 decisions) and Yang et al. [86] (1 file, 23 decisions) for eICU-CRD. This analysis identified 264 MIMIC-III and 57 eICU-CRD implementation decisions within the domains D1–D6 across 80 and 17 source files respectively, revealing substantial methodological heterogeneity unreported in the associated publications. Complete technical documentation is provided in Supplement S2.

Implementation dependencies exist among MIMIC-III repositories: For instance, Komorowski et al. [63] builds upon Johnson et al. [56], which derives from MIT-LCP. This dependency structure suggests potential propagation of methodological decisions, whether intentional design choices or incidental defaults, across a substantial portion of published sepsis research. The two eICU-CRD repositories were independently developed with no direct code dependencies.

For parameter coverage (D1), analysis across MIMIC-III repositories identified 149 unique ITEMIDs with variation in cardiovascular monitoring (18–28 ITEMIDs). Total parameter coverage ranged from 125 ITEMIDs (Komorowski et al. [63]) to 143 ITEMIDs (Johnson et al. [28]). Temporal window definitions (D2) varied substantially. Rosnati et al. [45] calculated SOFA scores relative to ICU admission with hourly resolution, while Johnson et al. [56] and Komorowski et al. [63] calculated within infection-relative windows (48-hour retrospective, 24-hour prospective around suspected infection onset). Missing data strategies (D4) showed the highest decision complexity (8–16 decisions per MIMIC-III repository). Johnson et al. [56] and MIT-LCP employed zero-imputation as primary strategy, reducing SOFA scores when measurements were absent. In contrast, Komorowski et al. [63] implemented multi-stage imputation procedures combining linear interpolation, k-nearest neighbor imputation, and variable-specific defaults. For SOFA calculation (D5), baseline definition varied fundamentally. Johnson et al. [56], MIT-LCP, Komorowski et al. [63], and Chen et al. [92] defined baseline as zero (interpreting the definition as absolute SOFA ≥ 2), while Rosnati et al. [45] implemented dynamic baseline using the lowest observed SOFA. For infection detection methods (D6), all four MIMIC-III repositories implemented antibiotic-culture temporal matching criteria [10]. Neither eICU-CRD repository implemented antibiotic-culture matching, both relied on APACHE related diagnosis codes. Complete domain-by-domain analysis is provided in Supplement S2.

The practical impact is illustrated by MIMIC-III *MetaVision* cohorts. Johnson et al. [56] reported 49.1 % detection rate, while overlapping implementations using the same source population reported 8.8 % – 42.0 % (Table 1). In eICU-CRD, differing temporal windows (24h vs. 72h), parameter coverage (32 vs. 14 identifiers), ICU-LOS requirements (≥ 72h vs. > 24h), and infection criteria (APACHE and ICD vs. APACHE only) would produce different cohorts from identical source data.

## Discussion

This systematic review of 44 MIMIC-III and 20 eICU-CRD studies reveals that the current sepsis definition, despite providing a standardized clinical framework, results in considerable variability in sepsis detection rates. Sepsis detection rates ranged from 3.4 % to 65.2 % in MIMIC-III (median 26.4 %) and from 9.8 % to 47.9 % in eICU-CRD (median 18.7 %); between-study differences, not sampling error, accounted for almost all of this variability (with such large cohorts, sampling error is negligible), indicating divergences in computational implementation decisions. The unit of analysis (patients, admissions or ICU stays) is itself a source of rate variation, since it mechanically changes the denominator: admission- and ICU-stay-based cohorts showed lower detection rates than patient-based cohorts, although no study characteristic reached significance in the meta-regression. Documentation rates were low across all domains examined, and source code analysis of six major repositories identified 321 specific implementation decisions that would produce different patient cohorts from identical data. The observed variability stems from three interconnected mechanisms rather than random methodological choices: First, publicly available ICU databases present data quality challenges requiring numerous undocumented decisions. MIMIC-III contains over 12,000 unique parameter names (e.g., > 100 names for ventilation parameters), physiologically implausible outliers (e.g., mean arterial pressure: 989999 mmHg), and dual information systems with different documentation practices (*CareVue* 2001–2008, *MetaVision* 2008–2012). These challenges are not database-specific; systematic comparison across five major ICU databases confirms that inconsistent diagnostic coding, variable temporal resolution, and heterogeneous parameter naming represent barriers to harmonization [93].

Second, missing data handling for SOFA components varies across studies. Our source code analysis revealed diverse approaches including zero-imputation (assuming normal organ function), multi-stage imputation procedures combining interpolation with k-nearest neighbor methods, and variable-specific defaults. Zero-imputation, while computationally simple, may underestimate dysfunction in critically ill patients where missing measurements may indicate deterioration rather than normality. Infection detection similarly requires choosing between antibiotic-culture temporal matching [10] (temporal precision but limited to treated infections) and ICD-9 diagnostic codes (broader coverage but dependent on variable coding practices).

Third, implementation choices may be shared across research groups. The similar median detection rates between Set A and Set B studies (24.9 % vs. 27.9 %) suggest that within-study methodological choices have greater impact than between-study patient population differences.

Our analysis further reveals patterns consistent with shared computational approaches across studies. In the eICU-CRD dataset, we observed two distinct clusters of studies reporting identical detection rates and cohort sizes.

Cluster 1 (34.94 % detection rate): Three publications by Zhang and colleagues [73–75] report identical cohorts (48,694 septic cases from 139,367 patients) while investigating different research questions.

Cluster 2 (16.60 % detection rate): Four publications—Lin et al. [82, 83], Shen et al. [84], and Chang et al. [85]—report identical cohorts (23,136 septic cases from 139,367 patients) across three geographically distinct institutions. Author analysis reveals a common co-author affiliated with a professional data analytics service across three of four publications [82, 83, 85]. This suggests that professional data analytics services may propagate specific implementation decisions across independent research groups without shared code documentation.

Detection rates showed no trend toward convergence over time (Figure 2), suggesting that awareness of implementation heterogeneity alone does not drive methodological consensus. These findings have implications for future sepsis definitions.

The recent introduction of SOFA-2 [94], with additional parameters and updated scoring thresholds, will likely increase the implementation heterogeneity documented here. Additional complexity of SOFA-2 introduces new decision points across all six methodological domains (D1–D6), from defining “missing” versus “not clinically indicated” parameters to handling selective clinical application of contemporary variables and accommodating variable temporal resolution across monitoring devices. To prevent these challenges repeating, we recommend that SOFA-2 deployment include proactive standardization: reference implementations with comprehensive D1–D6 documentation released simultaneously with clinical definitions, standardized test datasets for validation, mandatory implementation reporting integrated into existing guidelines (TRIPOD [19], CONSORT [95], STROBE [17]), and community-driven consensus on edge case handling before widespread adoption.

This study provides the first systematic multi-level analysis combining main text and source code examination to characterize variability in sepsis case detection. The six-domain framework (D1–D6) offers a structured approach for assessing and reporting methodological decisions. Our analysis of 321 implementation decisions across six major repositories quantifies previously undocumented sources of heterogeneity. Unlike studies documenting heterogeneity [9, 96, 97], our approach systematically characterizes the specific implementation decisions contributing to this variability.

Several limitations warrant consideration. First, our inclusion criteria restricted the corpus to open access publications, introducing a potential selection bias. Second, our analysis focused on MIMIC-III and eICU-CRD; generalizability to other datasets (MIMIC-IV, AmsterdamUMCdb, institutional databases) requires further research. Third, although a meta-regression on study-level characteristics explained little of the variability (about 9 % of the between-study variance; residual *τ*^2^ = 0.72), the low documentation rates (> 65 % undocumented for most domains) precluded regression quantifying the contribution of individual D2–D6 implementation decisions, and the extreme residual heterogeneity limited the power to resolve individual moderators. Fourth, we focused on methodological heterogeneity without clinical outcome validation; prospective evaluation using expert-adjudicated ground truth labels would be valuable to determine whether higher or lower detection rates better capture clinically meaningful sepsis.

The observed heterogeneity has potential implications for sepsis research reproducibility. That algorithmic implementation details, rather than the conceptual Sepsis-3 framework, drive this variability challenges the assumption that standardized definitions lead to comparable results across implementations. The predominance of undocumented methodological choices (> 65 % for most domains) points to a documentation gap that limits validation, comparison, and clinical implementation of complex multi-stage data processing methods. Comparable documentation gaps have been identified across clinical prediction modeling more broadly, with 82 % of 152 studies failing to report sample size calculations and 95 % omitting calibration metrics [98]. These challenges likely extend beyond the Sepsis-3 framework to other disease phenotypes derived from electronic health records that require complex operationalization from clinical data [99]. For the many sepsis studies employing machine learning, this is particularly consequential: prediction models trained on differently labeled cohorts may achieve comparable performance metrics while predicting distinct clinical phenotypes, compromising cross-study comparability and clinical translation.

The 321 implementation decision points identified provide a foundation for targeted standardization efforts. Therefore, we recommend: (1) mandatory reporting of implementation details across D1–D6 domains as supplementary material (a proposed reporting checklist is provided in Supplement S3), (2) publication of version-controlled source code, (3) development of consensus reference implementations for common databases, and (4) integration of sepsis label calculation standards into existing reporting guidelines, analogous to CONSORT for clinical trials, TRIPOD for prediction models, and STROBE for observational studies. Without such standardization, the clinical utility of sepsis prediction models will remain limited by implementation variability rather than model performance, with direct implications for the development of reliable clinical decision support systems.

### Conclusion

This systematic review shows that sepsis case detection varies substantially across studies, despite using the same sepsis definition, source databases and cohort specifications. We demonstrated that such heterogeneity is attributed to poor methodological documentation across six key domains: parameter coverage, temporal windows, aggregation strategies, missing value handling, SOFA calculation and infection detection methods. We recommend standardized reporting of sepsis case definitions and publication of version-controlled source code to improve reproducibility and robustness of sepsis prediction models.

## Supporting information

Supplement S1: Search Strategy, Dataset Selection, and Statistical Analysis

Supplement S2 - Systematic Source Code Analysis

Supplement S3 - Reporting Checklist for Automated Sepsis Case Detection

## Data Availability

All data supporting the findings of this study are available in a Zenodo repository (https://doi.org/10.5281/zenodo.18612350). The repository contains literature search exports, PRISMA screening records, detection rate and methodology classification data, source code analysis results, and Python & R scripts for reproducing all figures. Raw screening spreadsheets are available from the corresponding author upon reasonable request.

