## Supplement S1: Search Strategy, Dataset Selection, and Statistical Analysis for "Variability in Automated Sepsis Case Detection: A Systematic Analysis of Implementation Methods in Clinical Data Repositories"

### Supplement S1: Search Strategy and Dataset Selection

#### 1. Rationale for Dataset Selection

Based on institutional experience and initial literature review, we identified four potentially relevant, frequently-used publicly accessible intensive care databases: MIMIC-III [1], MIMIC-IV [2], eICU [3] (US-based), and AmsterdamUMCdb [4] (European). Dataset selection prioritized sufficient publication volume for systematic analysis, healthcare system comparability to control for system-level confounders, and institutional independence to avoid patient overlap.

We evaluated four major publicly available intensive care databases. MIMIC-III (Medical Information Mart for Intensive Care III) contains data from 46,520 patients admitted to Beth Israel Deaconess Medical Center between 2001 and 2012. MIMIC-IV extends coverage from the same institution through 2008–2019 with 299,712 patients. The eICU Collaborative Research Database aggregates data from 208 US hospitals covering 2014–2015 with 200,859 patients. AmsterdamUMCdb contains de-identified health records from over 20,000 ICU admissions at Amsterdam UMC from 2003–2016. Systematic literature searches (detailed in Section 2) quantified the Sepsis-3 [5] publication volume for each database: MIMIC-III yielded 278 publications (9.2 % of 3,020 total), MIMIC-IV produced 264 (22.6 % of 1,168), eICU contributed 110 (15.7 % of 700), and AmsterdamUMCdb yielded 17 (19.1 % of 89).

MIMIC-III was selected as the primary dataset based on the largest publication corpus (278 Sepsis-3 implementations), the longest publication history with extensive community knowledge and validated implementations, and robust technical documentation. eICU was selected as the validation dataset: as a multi-center database spanning 208 hospitals, it provides independent validation while maintaining the same US healthcare context, controlling for system-level confounders in documentation practices and clinical workflows.

MIMIC-IV was excluded because both MIMIC-III and MIMIC-IV originate from the same institution with overlapping time periods (2001–2012 vs. 2008–2019), creating substantial patient overlap that precludes independent validation. AmsterdamUMCdb was excluded due to insufficient publication volume (17 Sepsis-3 publications) and differences in healthcare system context (European vs. US) that would introduce confounding variation in clinical documentation practices and data capture workflows.

#### 2. Search Strategy

We conducted systematic literature searches in PubMed and Web of Science covering publications from January 1, 2016 through December 31, 2024. This timeframe begins with the Sepsis-3 definition publication and extends through the most recent available literature. The search strategy employed two complementary approaches: terms-based searches identifying publications mentioning specific terms in titles or abstracts, and citation-based searches identifying publications citing the original dataset or Sepsis-3 definition papers. Table 1 summarizes all search queries.

**Table 1.** Search queries by dataset. All searches were restricted to publications from January 1, 2016 through December 31, 2024. PubMed queries searched title and abstract fields; Web of Science queries employed equivalent syntax with TI and AB field tags. Citation searches identified publications citing the respective source paper.

| Target | Terms Search (PubMed syntax) | Citation Source |
|---|---|---|
| Sepsis-3 | `sepsis-3 OR "sepsis 3" OR sepsis-iii OR "sepsis iii"` | Singer et al., JAMA 2016; 10.1001/jama.2016.0287 |
| MIMIC-III | `mimic-iii OR "mimic iii" OR "mimic-3" OR mimic3 OR "mimic 3"` | Johnson et al., Sci Data 2016; 10.1038/sdata.2016.35 |
| eICU | `eICU` | Pollard et al., Sci Data 2018; 10.1038/sdata.2018.178 |
| MIMIC-IV | `mimic-iv OR "mimic iv" OR "mimic-4" OR mimic4 OR "mimic 4"` | Johnson et al., Sci Data 2023; 10.1038/s41597-022-01899-x |
| Amsterdam-UMCdb | `"Amsterdam UMC db" OR AUMCdb OR AmsterdamUMCdb` | Thoral et al., Crit Care Med 2021; 10.1097/CCM.0000000000004916 |

Search results were exported as RIS files from Web of Science or converted from TXT to RIS format from PubMed. Duplicate entries were removed within and across databases. Intersection analysis identified publications appearing in both Sepsis-3 and dataset-specific searches, and manual verification confirmed unique DOIs and publication metadata.

#### 3. Search Results

Table 2 presents the overall publication counts by search type for each dataset. Table 3 details the source-specific intersections for the two datasets included in the main analysis.

**Table 2.** Publication counts by search type for each ICU dataset and Sepsis-3. Combined counts reflect deduplicated totals across both search strategies. Intersection with Sepsis-3 indicates publications appearing in both the dataset-specific and Sepsis-3 corpora.

| Dataset | Terms | Citations | Combined | Intersection with Sepsis-3 |
|---|---|---|---|---|
| MIMIC-III | 1,279 | 2,541 | 3,020 | 278 (9.2 % of MIMIC-III corpus) |
| MIMIC-IV | 1,004 | 405 | 1,168 | 264 (22.6 % of MIMIC-IV corpus) |
| eICU | 413 | 559 | 700 | 110 (15.7 % of eICU corpus) |
| AmsterdamUMCdb | 20 | 84 | 89 | 17 (19.1 % of AmsterdamUMCdb corpus) |
| Sepsis-3 | 835 | 11,558 | 11,778 | — |

**Table 3.** Sepsis-3 intersections by literature database source for MIMIC-III and eICU. Additional references were identified through manual screening of seminal articles. Total screened represents the final number of unique publications entering the PRISMA screening process.

| Dataset | PubMed ∩ S-3 | WoS ∩ S-3 | Combined | Add. refs. | Total |
|---|---|---|---|---|---|
| MIMIC-III | 115 | 262 | 278 | 8 | 286 |
| eICU | 47 | 103 | 110 | 0 | 110 |
| Total | 162 | 365 | 388 | 8 | 396 |

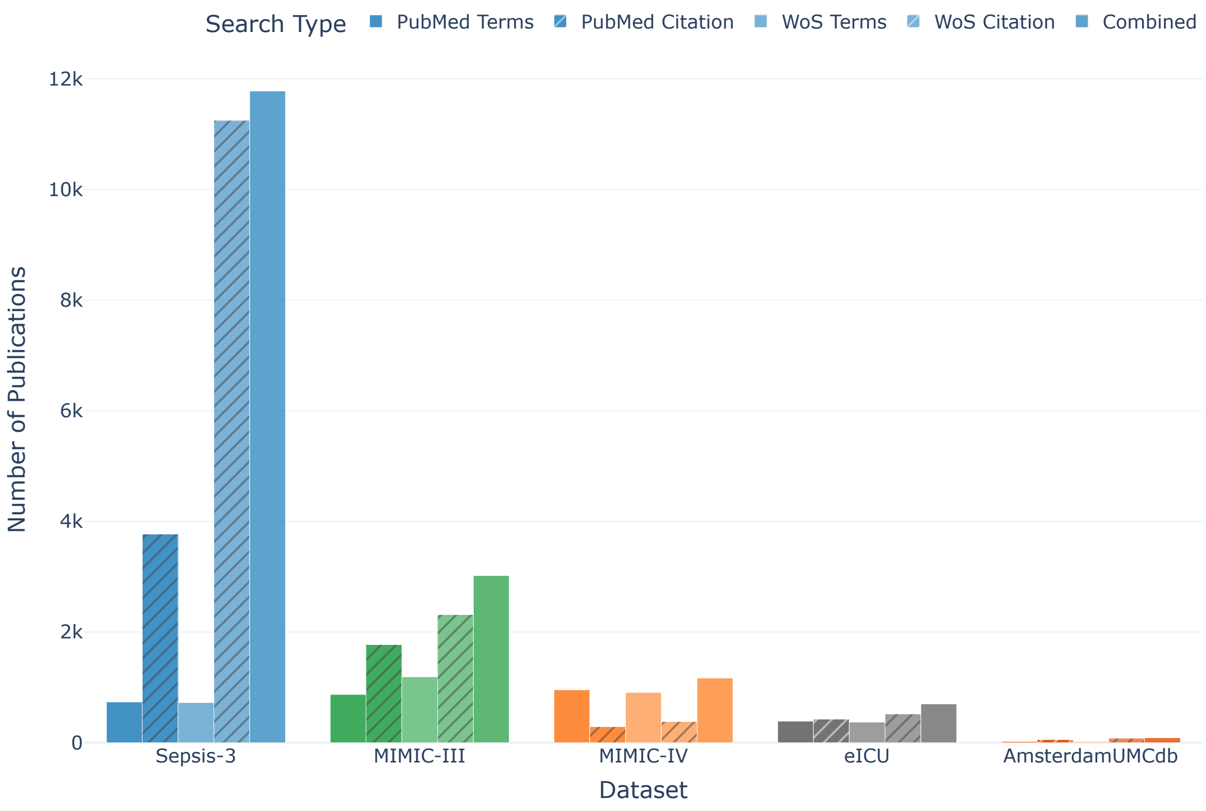


**Figure 1.** Total publications identified per dataset and search type (terms-based vs. citation-based) from 2016–2024.

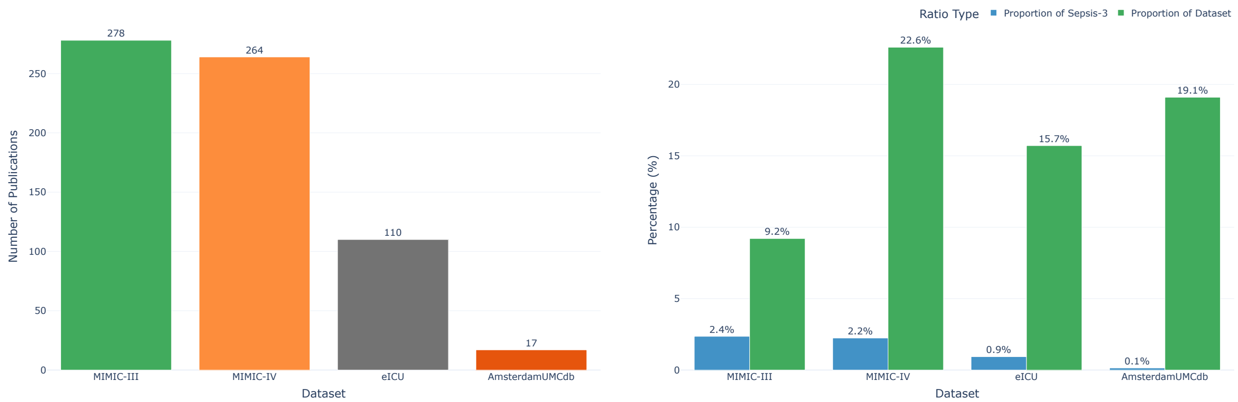


**(a)** Absolute counts

**(b)** Proportional representation

**Figure 2.** Sepsis-3 intersections by dataset. (a) Number of publications at the intersection of Sepsis-3 and each ICU dataset. MIMIC-III shows the largest absolute corpus ($n = 278$), followed by MIMIC-IV ($n = 264$), eICU ($n = 110$), and AmsterdamUMCdb ($n = 18$). (b) Proportion of Sepsis-3 publications relative to total dataset publications. MIMIC-IV shows the highest proportion (22.6 %), followed by AmsterdamUMCdb (20.0 %), eICU (15.7 %), and MIMIC-III (9.2 %).

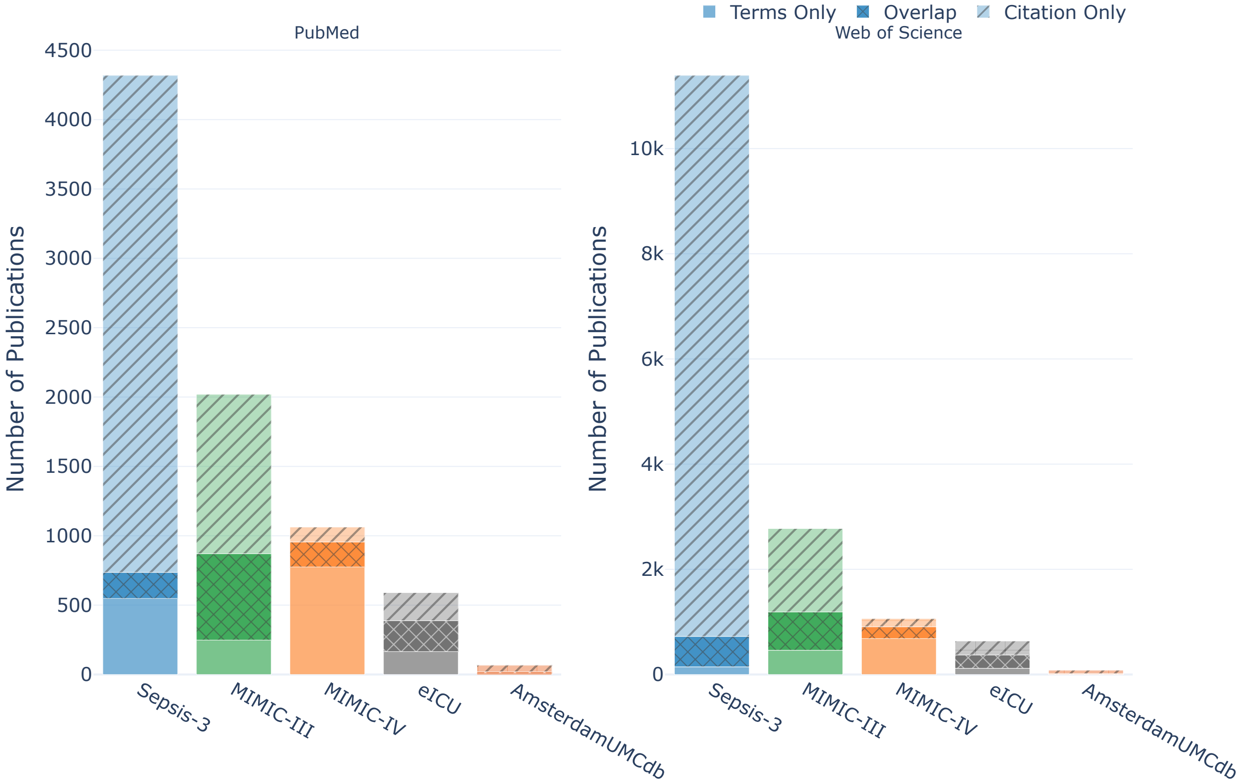


**Figure 3.** Overlap between terms-based and citation-based searches for each dataset. The limited overlap highlights the necessity of using both search strategies to achieve comprehensive coverage.

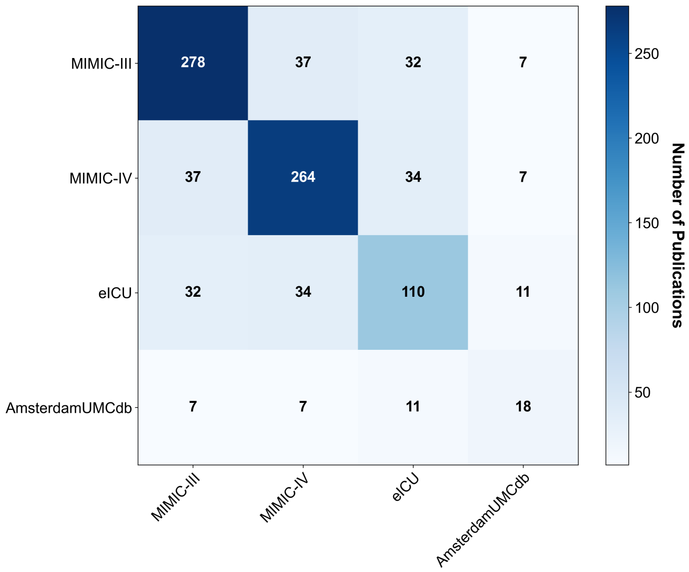


**Figure 4.** Overlaps between datasets within the Sepsis-3 publication corpus. Minimal overlap between MIMIC-III and eICU confirms institutional independence.

#### 4. Initial Screening Results

From the 278 MIMIC-III publications at the intersection with Sepsis-3, we applied inclusion and exclusion criteria following PRISMA guidelines. Inclusion criteria required English-language publications from January 2016 through December 2024 with open access full text, explicit MIMIC-III and Sepsis-3 implementation, and sufficient methodological details for sepsis label calculation. Exclusion criteria eliminated review articles without original analysis, conference abstracts without full papers, studies mentioning but not implementing Sepsis-3, and studies with insufficient methodological documentation. Title and abstract screening reduced the initial pool to 80 candidates; full-text assessment further refined the corpus to 44 publications included for detailed analysis.

For eICU, identical criteria were applied to the 110 publications at the intersection with Sepsis-3. Title and abstract screening reduced the pool to 38 candidates; full-text assessment refined the corpus to 20 publications included for detailed analysis.

#### 5. Examples of Screening and Eligibility Criteria

The screening and eligibility criteria are defined in the Methods and summarised in the PRISMA flow diagram. The accessibility, dataset, and Sepsis-3 criteria were straightforward to apply; the remaining four concern how a study derived and reported its cohort and detection rate, which we illustrate with examples below.

A study provided *label calculation details* when, beyond applying the Sepsis-3 definition, it described at least one concrete step in the label calculation (e.g., parameter aggregation, the observation window, imputation, or outlier handling), the infection-identification method, or a link to source code. Johnson et al. [6], Rosnati et al. [7], and Komorowski et al. [8] gave such details, backed by code. Studies that gave neither, such as those reusing the precomputed PhysioNet/Computing in Cardiology Challenge labels [9] without their own derivation [10], were excluded.

A study's detection rate was *traceable* when a septic count and cohort size for the general adult cohort could be obtained in a consistent unit (patients, admissions, or ICU stays), whether reported or reconstructed by tracing the cohort-derivation path. Where only the cohort size was missing, we set it to the dataset's documented adult population, making the reconstructed rate a conservative lower bound; a missing septic count, by contrast, could not be substituted. Rosnati et al. [7] were included on this basis: they reported a septic count but only an all-ages starting cohort, not the age-restricted adult denominator matching the count, so we used the documented adult population. In contrast, Lin et al. [11] were excluded because they reported a single figure of 38,600 “adult sepsis patients” with no separate total cohort; because this figure coincides with the documented adult MIMIC-III population, it appears to denote the whole cohort rather than a detected sepsis subset, so no septic count could be placed over a denominator and no detection rate could be obtained or reconstructed.

A study showed *hidden conditional filtering* when its cohort was restricted to a clinical subgroup (a specific disease, organ dysfunction, or comparable clinical condition) before sepsis identification, leaving only a subgroup rather than a general adult sepsis rate. Peng et al. [12] were included because they reported a general adult Sepsis-3 cohort before narrowing to sepsis-associated acute kidney injury (SA-AKI), so a baseline detection rate could still be recovered. In contrast, Libório et al. [13] were excluded because their cohort was restricted to a specific clinical subgroup—critically ill patients with a positive fluid balance—before sepsis identification, leaving only a subgroup rather than a general adult sepsis rate.

A study had *incomparable sepsis labels* when it did not provide a single sepsis cohort directly comparable with the others, either because it reported several non-comparable rates without an objective basis for selecting one, or because its cohort or data segment differed systematically from the standard reference population. Cohen et al. [14] were excluded because they reported multiple sepsis cohort sizes for the same MIMIC-III population (867–2,178 admissions) depending on the onset definition, with no objective basis for selecting a single estimate for comparison.

#### 6. Statistical Analysis of Detection-Rate Variability

We summarised the detection rates with a one-stage generalized linear mixed model (GLMM), fitted to the 58 studies with both a septic count and a cohort size: a binomial–normal random-effects meta-analysis with a study-level random intercept on the logit scale. We preferred this one-stage model to the two-stage DerSimonian–Laird method, whose moment estimator underestimates the between-study variance ($\tau^2$) when heterogeneity is high, as it is here; the DerSimonian–Laird analysis is reported as a sensitivity check. Rates were also compared across study characteristics using nonparametric tests on all 64 studies and a meta-regression on the 58. Each rate derives from a large cohort (about 12,000 to 200,000 patients or stays), so the studies are individually precise and Cochran's $Q$ and $I^2$ approach their maxima by construction. Inference was therefore based on $\tau^2$, the variance between studies on the logit scale, together with the observed range of rates.

Detection rates ranged from 3.4 % to 65.2 %. In the GLMM, combining the studies gave a pooled rate of 22.9 % (95 % CI, 19.1 % to 27.2 %), but heterogeneity dominated this estimate: with $\tau^2 = 0.79$ (95 % CI, 0.56 to 1.17), the 95 % prediction interval for a new study extended from 5 % to 63 %. The same pattern held within each database and in the robustness subsets. The two-stage DerSimonian–Laird analysis gave the same pooled rate but a smaller between-study variance ($\tau^2 = 0.46$), illustrating its downward bias at high heterogeneity (Table 4).

**Table 4.** Heterogeneity of the detection rates. The primary analysis is the one-stage binomial–normal GLMM, computed for each database and for both combined (all 58 studies with both counts); it is refitted to two robustness subsets (reported denominators only; patients only), and the two-stage DerSimonian–Laird estimate is shown as a sensitivity check. $k$ is the number of studies, $I^2$ the proportion of variation due to differences between studies, and $\tau^2$ the variance between studies on the logit scale. The $I^2$ values are computed from Cochran's $Q$ (not the GLMM) and are near their maximum in every row because each cohort is very large, so inference was based on $\tau^2$.

| | | $k$ | $I^2$ | $\tau^2$ |
|---|---|---|---|---|
| Main analysis (GLMM) | MIMIC-III | 38 | 99.98 % | 1.05 |
| | eICU-CRD | 20 | 99.99 % | 0.29 |
| | Both databases | 58 | 99.98 % | 0.79 |
| Robustness (GLMM) | Reported denominators only | 33 | 99.98 % | 0.95 |
| | Patients only | 29 | 99.98 % | 0.62 |
| Sensitivity | DerSimonian–Laird (both databases) | 58 | 99.98 % | 0.46 |

No characteristic distinguished the rates in the nonparametric tests (Table 5). The meta-regression explained little of the variance ($\tau^2$ from 0.79 to 0.72, about 9 %), and no characteristic reached significance; the largest difference was for the unit of analysis, with cohorts of admissions or ICU stays showing lower rates than cohorts of patients (odds ratios about 0.6, $p \geq 0.08$). Any such tendency reflects the changing denominator rather than a real difference, consistent with the non-significant nonparametric test for the unit of analysis ($p = 0.25$).

**Table 5.** Association of each study characteristic with the detection rate. The column $p^{\dagger}$ gives a nonparametric test across all 64 studies (Wilcoxon rank-sum for characteristics with two levels, Kruskal–Wallis otherwise). The OR and final $p$ columns give the primary GLMM meta-regression across the 58 studies, with the odds ratio of each level relative to its reference. The main model contains database, entity (the unit of analysis), and cohort set; the remaining characteristics were each tested by adding them to this model individually.

| Characteristic | $p^{\dagger}$ | Level | Reference | GLMM OR (95 % CI) | $p$ |
|---|---|---|---|---|---|
| Database | 0.39 | eICU-CRD | MIMIC-III | 0.83 (0.47–1.46) | 0.52 |
| Entity | 0.25 | Admissions | Patients | 0.56 (0.29–1.08) | 0.08 |
| | | ICU stays | Patients | 0.63 (0.35–1.12) | 0.11 |
| | | MetaVision | Patients | 0.60 (0.28–1.26) | 0.18 |
| Cohort set | 0.31 | Set B | Set A | 1.25 (0.71–2.18) | 0.44 |
| Study type | 0.17 | *added exploratorily; no significant coefficient* | | | |
| Infection detection method | 0.57 | *added exploratorily; no significant coefficient* | | | |
| Code availability | 0.70 | *added exploratorily; no significant coefficient* | | | |

Adding study type, infection detection method, or code availability to the main model individually did not improve the fit or produce a significant coefficient. The variance between studies remained essentially unchanged (residual $\tau^2$ 0.67 to 0.72 versus 0.72 for the main model), so these characteristics had little power to explain the heterogeneity.
