## Supplement S2 - Systematic Source Code Analysis for "Variability in Automated Sepsis Case Detection: A Systematic Analysis of Implementation Methods in Clinical Data Repositories"

### 1. Introduction

The Sepsis-3 definition [1] requires two conditions for sepsis classification: suspected infection (operationalized as temporal co-occurrence of antibiotic administration and body fluid culture [2]) and acute organ dysfunction (Sequential Organ Failure Assessment [SOFA] score $\geq 2$ points above baseline). While these clinical criteria appear straightforward, their computational implementation requires hundreds of technical decisions — from selecting database-specific parameter identifiers to choosing imputation strategies for missing values. These implementation decisions are rarely reported in publications, yet they directly affect which patients are classified as septic.

To systematically characterize implementation variability, we conducted a detailed source code analysis of six publicly available Sepsis-3 implementation repositories — four for MIMIC-III [3] and two for eICU-CRD [4]. This supplement documents the analysis methodology, the domain classification framework developed for organizing implementation decisions, and the complete cross-repository comparison results.

### 2. Methods

The source code analysis followed a structured three-phase approach (Table 1). During Phase 1, each source file was classified by relevance to the Sepsis-3 implementation into primary, supporting, or peripheral categories (Table 2).

Implementation decisions were classified into six domains (D1–D6) spanning the complete Sepsis-3 computational pipeline from raw clinical data to final label assignment (Table 3). Additionally, cohort selection decisions (age restrictions, information system constraints, multiple admission handling) were documented separately as study design choices outside the D1–D6 framework.

Each implementation decision was additionally classified by its position in the data processing pipeline (Table 4). This three-level scheme corresponds to the sequential stages of the Sepsis-3 computation pipeline: data extraction (Data-level), score computation (Score-level), label assignment (Label-

**Table 1.** Three-phase analysis approach for systematic source code review.

| Phase | Name | Description | Output |
|---|---|---|---|
| Phase 1 | Repository Profiling | Repository structure analysis: languages, dependencies, file organization, database platform, and commit history | Repository profile with file relevance classification |
| Phase 2 | Structured Extraction | Line-by-line analysis of all primary-relevance files: parameter identifiers, SQL logic, clinical thresholds, and temporal window definitions | Structured decision records with code references |
| Phase 3 | Cross-Repository Synthesis | Systematic comparison across repositories within each database, identifying consensus decisions, divergent implementations, and unique approaches | Comparative tables and decision matrices |

**Table 2.** File relevance classification categories.

| Category | Definition | Analysis Required |
|---|---|---|
| Primary | Directly implements SOFA scoring, infection identification, or Sepsis-3 label assignment | Full line-by-line extraction |
| Supporting | Provides helper functions, data transformations, or cohort definitions used by primary files | Targeted extraction of referenced components |
| Peripheral | Configuration, documentation, visualization, or unrelated analysis files | Review for context only |

level). Two independent reviewers classifying the same decision assign the impact level based on the pipeline stage at which the decision operates, ensuring reproducibility.

The following quality assurance measures were applied throughout the analysis: (1) All files classified as primary relevance were analyzed by two reviewers independently, with discrepancies resolved through discussion and consensus. (2) All extracted parameter mappings were validated against official database documentation — MIMIC-III ITEMIDs against the D_ITEMS table, eICU-CRD string identifiers against table schemas — to confirm parameter existence, correct labeling, and appropriate unit assignment. (3) After completing each file analysis, domain coverage was checked against a systematic checklist to ensure all SOFA organ systems, infection criteria, and temporal logic were documented where present. (4) Line number references were verified against specific repository commit hashes to ensure reproducibility, and analysis progress was tracked at the file level to ensure 100% coverage of primary relevance files. (5) Structured decision tables and repository profiles were synthesized from systematically extracted data using Claude Code (Anthropic Claude Opus 4.5, accessed January 2026) as a documentation assistant. All synthesized content was manually verified against original source code; the large language model was used solely for narrative synthesis and table formatting, while all implementation decisions, domain classifications, and impact assessments were determined through manual source code analysis.

**Table 3.** Domain classification framework for Sepsis-3 implementation decisions.

| | Domain | Description |
|---|---|---|
| D1 | Clinical Data Extraction & Parameter Identification | Selection of database-specific parameter identifiers (e.g., MIMIC-III ITEMIDs, eICU-CRD string identifiers), source table selection, unit conversion logic, and specimen type classification. These decisions determine which raw clinical measurements enter the SOFA calculation pipeline. |
| D2 | Temporal Window Definitions & Event Detection | Specification of time boundaries for data extraction, SOFA evaluation periods, and clinical event detection algorithms. A 24-hour rolling window with 1-hour stride produces hourly SOFA scores from which a ≥2-point increase can be detected; a single first-day window produces one SOFA score per ICU stay. This domain also encompasses vasopressor episode boundary detection (start time, end time, and continuation logic). |
| D3 | Parameter Aggregation Methods | Within any evaluation window, patients typically have multiple measurements for the same clinical parameter. The choice of aggregation function — minimum, maximum, or mean — determines which physiological state is represented in the SOFA score. Worst-case aggregation (e.g., minimum platelets, maximum bilirubin) produces systematically higher scores than mean-based approaches. |
| D4 | Missing Value Imputation Strategies | ICU patients have irregularly sampled measurements, and not all SOFA-relevant parameters are available for every patient at every time point. Whether treating absence as normal organ function (zero-imputation), carrying forward the last available measurement, or applying nearest-neighbor interpolation directly affects resulting SOFA scores and sepsis prevalence estimates. Decisions include carry-forward time limits (e.g., GCS carried forward up to 6 hours), component-specific defaults (e.g., GCS Motor = 6 for sedated patients), and assumed body weight when weight measurement is unavailable (e.g., 80 kg). |
| D5 | SOFA Score Calculation Variants | How clinical values are mapped to SOFA points according to organ-specific thresholds, how baseline organ function is defined for detecting acute deterioration, and how the final Sepsis-3 label is derived from the conjunction of SOFA increase and suspected infection. Key decisions include baseline SOFA definition (zero assumption vs. dynamic cumulative minimum), ventilation-dependent respiratory scoring thresholds, and weight-normalized vasopressor dosing for cardiovascular SOFA. |
| D6 | Infection Identification Methods | Under the Sepsis-3 definition, suspected infection — operationalized by Seymour et al. as temporal co-occurrence of antibiotic administration and body fluid culture — is a necessary condition for sepsis classification alongside SOFA ≥ 2. Key decisions include the primary methodological approach (antibiotic-culture temporal matching vs. ICD-9 diagnostic codes vs. Apache diagnosis fields), temporal window definitions for antibiotic-culture relationships (72 hours / 24 hours), and prescription-based vs. administration-based antibiotic identification. |

**Table 4.** Impact level classification.

| Impact Level | Pipeline Position | Definition | Criterion |
|---|---|---|---|
| Label-level | Final classification | Decisions that directly determine the Sepsis-3 label | Modification can directly reclassify patients |
| Score-level | Intermediate computation | Decisions affecting SOFA component or total score computation | Modification alters SOFA component scores |
| Data-level | Data extraction / preparation | Decisions affecting how clinical data is extracted | Modification changes the input data available |

### 3. Results

Six repositories were selected for detailed analysis — four implementing Sepsis-3 on MIMIC-III and two on eICU-CRD (Table 5). Among MIMIC-III repositories, AI_Clinician [6] builds upon sepsis3-mimic [5], which itself derives from mimic-code [7]; these dependencies suggest potential propagation of methodological decisions across implementations. The two eICU-CRD repositories are independent implementations with no direct code dependencies. Table 6 shows the domain distribution of the 321 identified D1–D6 implementation decisions. Additional cohort selection and other decisions are documented in the per-repository analysis files available in the Zenodo data repository.

**Table 5.** Repository overview.

| Repository | Database | Institution | Language | Files | D1–D6 | Publication |
|---|---|---|---|---|---|---|
| sepsis3-mimic | MIMIC-III | MIT-LCP | SQL (PgSQL) | 18 | 110 | [5] |
| AI_Clinician | MIMIC-III | Imperial Coll. London | MATLAB, Py | 3 | 36 | [6] |
| mimic-code | MIMIC-III | MIT-LCP | SQL (PgSQL, BQ) | 34 | 50 | [7] |
| MIMIC-III-sepsis-3-labels | MIMIC-III | Independent | Python, SQL | 25 | 68 | [8] |
| trajPLT | eICU-CRD | Independent | SQL (PgSQL) | 16 | 34 | [9] |
| AI-based-sepsis-enrichment | eICU-CRD | Independent | R, Python | 1 | 23 | [10] |

**Table 6.** Domain distribution of implementation decisions across repositories.

| Repository | Database | D1 | D2 | D3 | D4 | D5 | D6 | Total |
|---|---|---|---|---|---|---|---|---|
| sepsis3-mimic | MIMIC-III | 39 | 16 | 5 | 16 | 13 | 21 | 110 |
| AI_Clinician | MIMIC-III | 7 | 5 | 1 | 10 | 10 | 3 | 36 |
| mimic-code | MIMIC-III | 13 | 7 | 5 | 8 | 14 | 3 | 50 |
| MIMIC-III-sepsis-3-labels | MIMIC-III | 15 | 11 | 6 | 10 | 17 | 9 | 68 |
| *MIMIC-III subtotal* | | *74* | *39* | *17* | *44* | *54* | *36* | *264* |
| trajPLT | eICU-CRD | 11 | 5 | 4 | 5 | 6 | 3 | 34 |
| AI-based-sepsis-enrichment | eICU-CRD | 8 | 5 | 2 | 3 | 2 | 3 | 23 |
| *eICU-CRD subtotal* | | *19* | *10* | *6* | *8* | *8* | *6* | *57* |
| **Total** | | **93** | **49** | **23** | **52** | **62** | **42** | **321** |

**sepsis3-mimic** [5] (110 decisions). This implementation uses zero-imputation for missing SOFA components, treating absent values as normal organ function. SOFA scores are calculated relative to suspected infection onset using a 72-hour window (−48h to +24h). Infection identification relies on prescription-based (POE) antibiotics, capturing 2.6 times more suspected infections than IV-only approaches (i.e., identifying antibiotics solely from intravenous administration records). Notable technical decisions include an ML-based arterial specimen classification with a 75% confidence threshold and a MetaVision-only population constraint that excludes >60% of MIMIC-III ICU stays. Vasopressor calculations differ between information systems: CareVue rates require weight normalization while MetaVision rates are used directly.

**AI_Clinician** [6] (36 decisions). Designed for reinforcement learning, this implementation discretizes continuous clinical data into 4-hour blocks and employs a multi-stage imputation pipeline: linear interpolation for parameters with <5% missing values, followed by k-nearest neighbor imputation ($k=1$, standardized Euclidean distance) for remaining gaps. Vasopressor doses are standardized to norepinephrine equivalents assuming 80 kg body weight. Infection identification uses asymmetric temporal windows (24h antibiotic-first, 72h culture-first). The P/F ratio is recalculated after imputation, and baseline SOFA is explicitly set to zero in the source code.

**mimic-code** [7] (50 decisions). As the MIT-LCP reference implementation, this repository provides the foundation from which sepsis3-mimic and AI_Clinician derive. It uses a first-day temporal scope (±24h from ICU admission) with zero-imputation for missing values. Sedated patients are assigned GCS=15 following SAPS II methodology, and an 80 kg weight fallback is used for vasopressor dose normalization. Respiratory scoring applies ventilation-dependent thresholds with an 8-hour continuation window. Infection identification uses asymmetric windows (72h retrospective, 24h prospective). The implementation supports dual database platforms (PostgreSQL and BigQuery) and extracts laboratory values through pre-computed pivot tables.

**MIMIC-III-sepsis-3-labels** [8] (68 decisions). This repository provides the highest temporal resolution among the analyzed implementations, calculating SOFA scores hourly throughout the ICU stay using a 24-hour rolling maximum for component aggregation. Uniquely, it defines baseline SOFA as a dynamic cumulative minimum rather than assuming zero. Like sepsis3-mimic, it implements ML-based arterial specimen classification (75% confidence) and system-specific vasopressor calculations (CareVue weight-normalized vs. MetaVision direct rates). Additional distinctive features include GU irrigant volume correction for accurate urine output measurement and symmetric Seymour criteria (72h/24h) validated against 140 antibiotic patterns.

**trajPLT** [9] (34 decisions). This eICU-CRD implementation contains two parallel SOFA calculation methods (first_day_sofa and SOFA_OXY) that differ in time windows, cardiovascular scoring approach (binary vs. dose-dependent), and vasopressor coverage (4 vs. 7 agents). Infection is identified through Apache diagnosis categories and ICD codes rather than Seymour antibiotic-culture criteria. The implementation includes an SpO2/FiO2 fallback when arterial blood gas data is unavailable and uses dual vasopressor data sources (infusiondrug and medication tables with HICL codes). A study-specific ICU length of stay requirement of ≥72 hours is applied.

**AI-based-sepsis-enrichment-strategy** [10] (23 decisions). This implementation relies on external pre-calculated tables for SOFA scoring, rendering the underlying methodology invisible in the analyzed code. Infection identification uses the simplest approach among all repositories — a single string match against Apache admission diagnosis fields, without ICD codes or Seymour criteria. Temporal windows for laboratory data are notably wide (72h: −24h to +48h from ICU admission). The repository tracks 5 vasopressors (excluding dobutamine and milrinone), assumes 77 kg body weight when measurements are unavailable, and contains a high-severity bug in base excess validation that flips negative values to positive.

**MIMIC-III cross-repository comparison.** Table 7 compares SOFA component implementations across the four MIMIC-III repositories, and Table 8 summarizes ITEMID coverage. All four repositories implemented identical SOFA scoring thresholds per the original SOFA definition [1] (Respiratory: PaO2/FiO2 at 400, 300, 200, 100; Cardiovascular: dopamine >15 or epinephrine/norepinephrine >0.1; Neurological: GCS 13–14, 10–12, 6–9, <6; Renal: creatinine 1.2, 2.0, 3.5, 5.0; Hepatic: bilirubin 1.2, 2.0, 6.0, 12.0; Coagulation: platelets 150, 100, 50, 20), used the same core laboratory ITEMIDs for creatinine (50912), bilirubin (50885), and platelets (51265), and restricted cohorts to adult patients (≥16 or

≥18 years). Despite these consensus decisions, substantial variation was observed across multiple dimensions (Table 9).

**Table 7.** SOFA component implementation comparison across MIMIC-III repositories.

| Component | sepsis3-mimic | AI_Clinician | mimic-code | MIMIC-III-s3-labels |
|---|---|---|---|---|
| *Cardiovascular* | | | | |
| Vasopressor logic | CareVue weight-normalized, MetaVision direct rates | Norepinephrine equivalents, 80 kg fallback | Weight-normalized, 80 kg fallback | 2-month weight averaging + echo backup |
| Episode detection | Stop-signal detection, rate = 0 triggers new episode | 4-hour discretization blocks | Complex event aggregation | Stop-signal + ‘D/C’d’ detection |
| Weight strategy | System-dependent | 80 kg when missing | 80 kg when missing | 2 months before admission to discharge |
| *Respiratory* | | | | |
| Ventilation detection | Inherited from mimic-code | Binary detection, missing = non-ventilated | 3-stage event logic, 8-hour continuity | 3-stage + oxygen device detection |
| Event algorithm | 3-stage: initiate-continue-terminate | Simple binary assumption | 3-stage: initiate-continue-terminate | 3-stage + comprehensive termination |
| *Neurological* | | | | |
| GCS components | CareVue + MetaVision (all 3) | Motor component only | CareVue + MetaVision (all 3) | CareVue + MetaVision + 6-hour forward-fill |
| *Renal* | | | | |
| Criteria | Creatinine OR urine output | Realtime + preadmission measurements | Pivoted approach | GU irrigant volume correction |
| *Baseline Definition* | | | | |
| SOFA baseline | Zero-imputation (implicit zero) | Explicit zero assumption | Zero-imputation (implicit zero) | Dynamic cumulative minimum |

**Table 8.** ITEMID coverage by SOFA component across MIMIC-III repositories.

| SOFA Component | Total | sepsis3-mimic | AI_Clinician | mimic-code | MIMIC-III-s3-labels |
|---|---|---|---|---|---|
| Respiratory | 83 | 82 | 74 | 82 | 78 |
| Cardiovascular | 28 | 25 | 20 | 28 | 18 |
| Neurological | 6 | 6 | 2 | 6 | 6 |
| Renal | 30 | 26 | 27 | 2 | 27 |
| Hepatic | 1 | 1 | 1 | 1 | 1 |
| Coagulation | 1 | 1 | 1 | 1 | 1 |
| Total | 149 | 141 | 125 | 14 | 131 |

Notes: sepsis3-mimic inherits mechanical ventilation ITEMIDs from mimic-code rather than implementing independent ventilation detection logic. AI_Clinician implements simplified binary ventilation classification and documents only GCS Motor component ITEMIDs (454, 223901); Verbal and Eyes may be implemented but were not captured during extraction. AI_Clinician additionally separates urine output into realtime (24 ITEMIDs) and preadmission (2 ITEMIDs: 40060, 226633). mimic-code uses precomputed pivoted tables with label-based parameter selection; underlying ITEMIDs are equivalent but abstracted through the pivot table layer. The complete ITEMID cross-repository mapping (149 ITEMIDs) is provided in the Zenodo data repository (file: mimic_itemid_mapping.csv).

**Table 9.** Key implementation differences across MIMIC-III repositories.

| **Aspect** | **Implementation variation** |
|---|---|
| Baseline SOFA definition | Zero assumption (sepsis3-mimic, AI_Clinician, mimic-code) vs. dynamic cumulative minimum (MIMIC-III-sepsis-3-labels) |
| Temporal resolution | Hourly (MIMIC-III-sepsis-3-labels) vs. 4-hour blocks (AI_Clinician) vs. infection-window-based (sepsis3-mimic) vs. first-day (mimic-code) |
| Seymour criteria windows | Symmetric 72h/24h (sepsis3-mimic, MIMIC-III-sepsis-3-labels) vs. asymmetric matching (AI_Clinician, mimic-code) |
| Weight normalization | Fixed 80 kg (AI_Clinician, mimic-code) vs. 2-month averaging (MIMIC-III-sepsis-3-labels) vs. system-dependent (sepsis3-mimic) |
| GCS assessment | All 3 components (sepsis3-mimic, mimic-code, MIMIC-III-sepsis-3-labels) vs. Motor only (AI_Clinician) |
| Cardiovascular ITEMIDs | 18–28 (up to 1.6-fold variation); mimic-code uniquely includes milrinone |
| Missing value strategy | Zero-imputation (sepsis3-mimic, mimic-code) vs. k-nearest neighbor $k=1$ (AI_Clinician) vs. dynamic minimum baseline (MIMIC-III-sepsis-3-labels) |
| Arterial specimen handling | ML-based classification with 75% threshold (sepsis3-mimic, MIMIC-III-sepsis-3-labels) vs. not implemented (AI_Clinician, mimic-code) |
| Urine output correction | GU irrigant volume subtraction (MIMIC-III-sepsis-3-labels) vs. standard extraction (others) |

These findings carry implications for reproducibility. Core parameters for hepatic, coagulation, and renal SOFA use identical ITEMIDs across all repositories, providing a stable foundation. However, cardiovascular coverage varies most — the 1.6-fold difference (18–28 ITEMIDs) may affect cardiovascular SOFA scoring, particularly for patients receiving phenylephrine, vasopressin, or milrinone. Neurological assessment differs fundamentally, with AI_Clinician's use of GCS Motor alone representing a qualitatively different approach from the full 3-component GCS used by other repositories. Respiratory implementation is similarly complex: while ITEMID counts appear similar (74–82), event detection algorithms differ substantially, ranging from binary detection to 3-stage initiate-continue-terminate logic with 8-hour continuation windows.

**eICU-CRD cross-repository comparison.** The two eICU-CRD repositories use string-based parameter identification (drug/lab names) rather than numeric ITEMIDs, and neither supports the Seymour antibiotic-culture temporal matching approach due to limited culture data availability. Table 10 compares SOFA component implementations and Table 11 summarizes parameter coverage. Both repositories use Apache diagnosis-based identification rather than Seymour antibiotic-culture criteria, extract creatinine, bilirubin, and platelets from the eICU lab table, and limit cohorts to adult patients (≥18 years) with first ICU stay only. Despite these shared characteristics, implementation decisions diverge substantially (Table 12).

**Table 10.** SOFA component implementation comparison across eICU-CRD repositories.

| Component | trajPLT | AI-based-sepsis-enrichment |
|---|---|---|
| *Cardiovascular* | | |
| Vasopressor source | infusiondrug + medication (dual) | infusionDrug only (single) |
| Vasopressors tracked | 7 (incl. dobutamine, milrinone) | 5 (excl. dobutamine, milrinone) |
| HICL codes used | Yes (medication table) | No |
| Dose-dependent scoring | Yes (SOFA_OXY) / No (first_day_sofa) | External (invisible) |
| Weight assumption | 80 kg | 77 kg |
| *Respiratory* | | |
| P/F ratio source | lab table + respiratorycharting | External (pf table) |
| SpO2/FiO2 fallback | Yes (SOFA_OXY) | Unknown (external) |
| Ventilation detection | 42 treatment strings, 3 source tables | External (eicu_mv table) |
| *Neurological* | | |
| GCS source | physicalexam OR apacheapsvar | nurseCharting ('GCS Total') |
| GCS components | 3 components (eyes, verbal, motor) | Total score only |
| Sedation handling | meds=1 → GCS=15 (first_day_sofa) | Not specified |
| *Renal* | | |
| Urine output source | intakeoutput / apacheapsvar | External (eicu_uop table) |
| *SOFA Calculation* | | |
| Calculation method | Internal (visible, dual implementations) | External (invisible) |

**Table 11.** eICU-CRD parameter coverage by SOFA component.

| SOFA Component | Total | trajPLT | AI-based | Difference |
|---|---|---|---|---|
| Respiratory | 7 | 7 | 2* | 5 invisible (external tables) |
| Cardiovascular | 16 | 16 | 7 | 9 not covered (HICL codes, dobutamine, milrinone) |
| Neurological | 5 | 4 | 1 | Different sources (components vs. total) |
| Renal | 3 | 3 | 2* | 1 source not used |
| Hepatic | 1 | 1 | 1 | Identical |
| Coagulation | 1 | 1 | 1 | Identical |
| Total | 33 | 32 | 14* | 19 parameters fewer/invisible |

*AI-based uses external pre-calculated tables whose methodology is not visible in the analyzed code.

The complete eICU-CRD parameter identification mapping (36 identifiers: 33 SOFA parameters + 3 sepsis identification) is provided in the Zenodo data repository (file: eicu_parameter_mapping.csv).

**Table 12.** Key implementation differences across eICU-CRD repositories.

| Aspect | Implementation variation |
|---|---|
| Vasopressor coverage | 7 agents including dobutamine and milrinone (trajPLT) vs. 5 agents (AI-based) |
| Vasopressor identification | Dual-source with HICL codes (trajPLT) vs. single-source string matching (AI-based) |
| SOFA calculation | Internal and fully visible (trajPLT) vs. external pre-calculated tables (AI-based) |
| GCS source | Dual-source with 3 components (trajPLT) vs. single-source total score (AI-based) |
| ICU LOS requirement | ≥72h (trajPLT) vs. >24h (AI-based) |
| Laboratory temporal window | 24h (trajPLT) vs. 72h (AI-based) |
| Weight assumption | 80 kg (trajPLT) vs. 77 kg (AI-based) |
| ICD codes for sepsis | Apache + ICD-9/10 codes (trajPLT) vs. Apache string match only (AI-based) |

Each repository also contributes unique methodological approaches. trajPLT implements dual SOFA calculations with SpO2/FiO2 ratio fallback, HICL code-based vasopressor identification, and 42 treatment strings for mechanical ventilation detection. AI-based relies on external pre-calculated tables (rendering SOFA methodology non-reproducible), implements a norepinephrine-equivalent formula (norepi + epi + phenyl/10 + dopa/150 + vaso × 2.5), and contains a base excess validation bug that flips negative values to positive.

These findings highlight two key reproducibility concerns for eICU-CRD implementations. First, string-based parameter identification introduces variability absent from MIMIC-III's numeric ITEMIDs — trajPLT's use of HICL codes represents a more standardized approach than pure string matching. Second, external table dependencies fundamentally reduce transparency: AI-based depends on pre-calculated tables whose methodology cannot be verified from the analyzed code, making the SOFA calculation non-reproducible. The clinically significant difference in vasopressor coverage (exclusion of dobutamine and milrinone) may additionally affect cardiovascular SOFA scoring.
