## Supplement S3 - Reporting Checklist for Automated Sepsis Case Detection for "Variability in Automated Sepsis Case Detection: A Systematic Analysis of Implementation Methods in Clinical Data Repositories"

### Supplement S3:
### Reporting Checklist for Automated Sepsis Case Detection

#### Variability in Automated Sepsis Case Detection: A Systematic Analysis of Implementation Methods in Clinical Data Repositories

**Falk Meyer-Eschenbach, MSc**[1,2,5,✉], **René Schmiedler, MSc**[1], **Johannes v. Stoephasius, BSc**[5], **Camille Zhang, MSc**[2], **Luis Kronfli, MD**[1], **Nicolas Frey, MSc**[1], **Anatol-Fiete Näher, MD**[4], **Jacques Ehret, MSc**[2], **Johanna Nothacker, PhD**[2], **Christof von Kalle, MD**[2], **Severin Kohler, MSc**[6,7], **Elias Grünewald, PhD**[1], **Andreas Edel, MD**[3], **Oliver Kumpf, MD**[3], **Jon Barrenetxea, PhD**[1], **Felix Balzer, MD, PhD**[1]

[1]Institute of Medical Informatics, Charité – Universitätsmedizin Berlin, Germany
[2]Clinical Study Center, Berlin Institute of Health, Berlin, Germany
[3]Klinik für Anästhesiologie und Intensivmedizin (CCM | CVK), Charité – Universitätsmedizin Berlin, Germany
[4]Digital Global Public Health, Hasso Plattner Institute, University of Potsdam, Potsdam, Germany
[5]Institute of Informatics, Humboldt University, Berlin, Germany
[6]Institute of Informatics, Freie Universität Berlin, Berlin, Germany
[7]Health Data Science Unit, University Hospital Heidelberg, Heidelberg, Germany

#### Supplement S3:
#### Reporting Checklist for Automated Sepsis Case Detection

This checklist is derived from the six-domain methodological framework (D1–D6) identified through systematic review and source code analysis of 64 studies implementing the Sepsis-3 definition on MIMIC-III and eICU-CRD databases. It is intended for researchers implementing automated sepsis case detection from electronic health records and for reviewers evaluating such studies. The checklist complements existing reporting guidelines (TRIPOD, CONSORT, STROBE) by addressing the computational phenotyping decisions that these frameworks do not cover, specifically the data processing pipeline from raw electronic health record data to final disease labels.

For each item, document the specific implementation choice in the "Notes / Reference" column. Provide a source code repository URL and commit hash where applicable. For studies with published source code, most items can be extracted directly from the codebase, substantially reducing the reporting effort.

##### Study Metadata & Cohort Definition

| ID | Reporting Item | Specification Required | Reported | Notes / Reference |
|---|---|---|---|---|
| S0-01 | Database and version | Name, version number, access date | ☐ | |
| S0-02 | Cohort source table | Primary table(s) used for patient selection | ☐ | |
| S0-03 | Age restriction | Minimum/maximum age thresholds applied | ☐ | |
| S0-04 | Multiple admission handling | Policy for patients with multiple ICU stays (first, last, all, longest) | ☐ | |

| ID | Reporting Item | Specification Required | Reported | Notes / Reference |
|---|---|---|---|---|
| S0-05 | Additional exclusion criteria | ICU length-of-stay requirements, missingness thresholds, outlier removal | ☐ | |
| S0-06 | Reported cohort sizes | Total patients/admissions screened and sepsis-positive count with detection rate | ☐ | |
| S0-07 | Source code availability | Repository URL and commit hash | ☐ | |
| S0-08 | Reference implementation | Upstream repository or prior implementation used as basis, if any | ☐ | |

**D1: Parameter Coverage**

| ID | Reporting Item | Specification Required | Reported | Notes / Reference |
|---|---|---|---|---|
| D1-01 | Respiratory parameters | PaO2, FiO2, SpO2 variants, mechanical ventilation identifiers, extubation and oxygen therapy parameters used to determine ventilation status | ☐ | |
| D1-02 | Cardiovascular parameters | Vasopressor names, MAP identifiers | ☐ | |
| D1-03 | Hepatic parameters | Bilirubin identifiers | ☐ | |
| D1-04 | Coagulation parameters | Platelet count identifiers | ☐ | |
| D1-05 | Renal parameters | Creatinine, urine output, and body weight identifiers | ☐ | |
| D1-06 | Neurological parameters | GCS component identifiers, sedation handling | ☐ | |
| D1-07 | Information system handling | How dual systems (e.g., CareVue/MetaVision in MIMIC-III) are handled: merged, separated, or single system only | ☐ | |
| D1-08 | Derived parameters | Whether any parameters are calculated from component values (e.g., MAP from SBP/DBP, total bilirubin from direct/indirect, PaO2/FiO2 ratio) and formulas used | ☐ | |

| ID | Reporting Item | Specification Required | Reported | Notes / Reference |
|---|---|---|---|---|
| D1-09 | Physiological range validation | Upper/lower bounds applied per parameter to exclude implausible values (e.g., MAP range, creatinine range, bilirubin range, temperature range) | ☐ | |
| D1-10 | Unit conversions | Conversions applied to raw data (e.g., temperature Fahrenheit to Celsius, weight lbs/oz to kg, bilirubin μmol/L to mg/dL) | ☐ | |
| D1-11 | Arterial specimen classification | How arterial vs. venous blood gas samples are distinguished for PaO2/FiO2 calculation (e.g., ML-based classification with probability threshold, specimen label, or no distinction) | ☐ | |
| D1-12 | Infection detection parameters | Antibiotic names/routes, culture types, or ICD code lists used | ☐ | |

**D2: Temporal Windows**

| ID | Reporting Item | Specification Required | Reported | Notes / Reference |
|---|---|---|---|---|
| D2-01 | SOFA calculation scope | Full ICU stay, fixed window around event, or rolling window | ☐ | |
| D2-02 | Aggregation window size | Duration of each assessment window (e.g., 1h, 6h, 24h) | ☐ | |
| D2-03 | Window step / offset | Sliding window step size, if applicable | ☐ | |
| D2-04 | Temporal reference point | ICU admission, infection onset, or other anchor point | ☐ | |
| D2-05 | Infection detection window | Temporal boundaries for antibiotic-culture matching (e.g., −48h to +24h relative to suspected infection onset) | ☐ | |

**D3: Aggregation Methods**

| ID | Reporting Item | Specification Required | Reported | Notes / Reference |
|---|---|---|---|---|
| D3-01 | Vital sign aggregation | Strategy for multiple measurements within window (worst-case, mean, median, most recent) | ☐ | |
| D3-02 | Laboratory value aggregation | Strategy for multiple lab results within window | ☐ | |
| D3-03 | Urine output calculation | Cumulative calculation method, rolling sum duration | ☐ | |
| D3-04 | Vasopressor aggregation | Dose aggregation for multiple concurrent vasopressors, norepinephrine-equivalent conversion, episode construction from discrete administrations (stop signal handling, gap threshold for new episodes, handling of missing stop signals) | ☐ | |
| D3-05 | Ventilation episode construction | Rules for determining continuous mechanical ventilation from discrete settings (e.g., gap threshold for episode termination, events that end ventilation such as extubation or oxygen therapy initiation, sequential settings treated as continuous) | ☐ | |
| D3-06 | GCS aggregation | Component-level vs. total score aggregation, handling of sedated patients | ☐ | |

**D4: Missing Data Handling**

| ID | Reporting Item | Specification Required | Reported | Notes / Reference |
|---|---|---|---|---|
| D4-01 | Primary imputation strategy | Method used when measurements are absent (zero-imputation, forward-fill, interpolation, kNN, multiple imputation) | ☐ | |
| D4-02 | Default assumption | Whether missing SOFA components default to normal (score = 0) or are handled differently | ☐ | |

| ID | Reporting Item | Specification Required | Reported | Notes / Reference |
|---|---|---|---|---|
| D4-03 | Component-specific handling | Any SOFA components with distinct imputation rules (e.g., GCS in sedated patients, urine output when not measured) | ☐ | |
| D4-04 | Default body weight | Assumed body weight when patient weight is unavailable, used for vasopressor dose normalization (e.g., 80 kg) | ☐ | |
| D4-05 | FiO2 default assumption | Value assumed when FiO2 is not recorded (e.g., room air at 21 %) | ☐ | |
| D4-06 | Carry-forward duration | Maximum time window for carrying forward previous measurements when current values are missing (e.g., GCS carry-forward up to 6 hours) | ☐ | |
| D4-07 | Minimum data requirements | Minimum number of measurements or time coverage required per patient/stay for inclusion | ☐ | |
| D4-08 | Distinction from prediction model preprocessing | Whether missing data handling for label calculation differs from preprocessing applied to prediction model features | ☐ | |

**D5: SOFA Calculation**

| ID | Reporting Item | Specification Required | Reported | Notes / Reference |
|---|---|---|---|---|
| D5-01 | SOFA threshold | Threshold used for sepsis determination (e.g., $\geq 2$ points) | ☐ | |
| D5-02 | Baseline SOFA definition | How baseline organ function is defined: assumed zero, admission SOFA, pre-ICU SOFA, or dynamic lowest-observed | ☐ | |
| D5-03 | Baseline determination window | Time window for establishing baseline SOFA, if not assumed zero | ☐ | |

| ID | Reporting Item | Specification Required | Reported | Notes / Reference |
|---|---|---|---|---|
| D5-04 | Component scoring table | Explicit scoring thresholds for each of the six SOFA components, including any modifications from the original definition | ☐ | |
| D5-05 | SpO2/FiO2 handling | Whether SpO2/FiO2 ratio is used as surrogate for PaO2/FiO2, and conversion method if applicable | ☐ | |
| D5-06 | Vasopressor dose thresholds | Dose cutoffs for cardiovascular SOFA scoring, norepinephrine-equivalent conversion factors for each vasopressor, unit conversions applied | ☐ | |

**D6: Infection Detection Methods**

| ID | Reporting Item | Specification Required | Reported | Notes / Reference |
|---|---|---|---|---|
| D6-01 | Primary method | Antibiotic-culture temporal matching, ICD codes, APACHE diagnosis codes, clinical documentation, or hybrid approach | ☐ | |
| D6-02 | Antibiotic criteria | Antibiotic list used, route restrictions (IV only vs. all routes), minimum duration requirements | ☐ | |
| D6-03 | Culture criteria | Culture types included (blood, urine, respiratory, other), timing relative to antibiotic administration | ☐ | |
| D6-04 | Temporal matching logic | Time window and directionality for antibiotic-culture co-occurrence (e.g., culture within ±24h of antibiotic, or antibiotic within 72h of culture) | ☐ | |
| D6-05 | ICD code list | Complete list of ICD-9/ICD-10 codes used for infection identification, if applicable | ☐ | |

| ID | Reporting Item | Specification Required | Reported | Notes / Reference |
|---|---|---|---|---|
| D6-06 | Suspected infection onset | How onset time is defined (first antibiotic, first culture, earliest of either) | ☐ | |

**Minimum Reporting Standard.** Based on the documentation gaps identified in our systematic review, we recommend that all studies implementing automated sepsis case detection report at minimum: (1) the complete specification for D5 (SOFA Calculation) and D6 (Infection Detection Methods), (2) the primary strategy for D4 (Missing Data Handling), (3) temporal window parameters from D2, and (4) a link to version-controlled source code. Full reporting across all domains (D1–D6) with accompanying code is strongly encouraged.
